# Challenges in Classification of *PKD1* Missense Variation in Autosomal Dominant Polycystic Kidney Disease

**DOI:** 10.64898/2026.07.30.26359376

**Authors:** Nicole Lehmann, Steven Koo, Yvonne Hort, Gopala Rangan, Gladys Ho, Rocio Rius, Amali Mallawaarachchi

## Abstract

**Purpose:** Autosomal Dominant Polycystic Kidney Disease is the most common monogenic kidney disease and largely due to variants in *PKD1*. We aimed to assess pathogenicity evidence for *PKD1* missense variants in disease databases and evaluate *in silico* pathogenicity prediction tool performance.

**Methods:** *PKD1* missense variants reported as pathogenic, likely pathogenic or likely benign were extracted from ClinVar and PKDB. Variants were re-classified using ACMG/AMP criteria to identify ‘truth sets’ of pathogenic and benign variants. *In silico* scores were obtained from five tools (SIFT, PolyPhen-2, CADD, REVEL, AlphaMissense) and evaluated using established thresholds. A Receiver Operating Characteristic curve analysis was performed using the *PKD1* variant truth sets.

**Results:** 346/389 (89%) reported disease-causing missense variants in *PKD1* were downgraded to Variants of Unknown Significance (VUS) using current classification criteria. Based on current thresholds, REVEL achieved the highest sensitivity of 62%, with specificity of 79%. AlphaMissense was the only tool not to misclassify any truth set variants, but many of the variant scores were between the pathogenic and benign thresholds.

**Conclusion:** A large majority of *PKD1* missense variants are classified as VUS with current pathogenicity criteria. Commonly used *in silico* tools, applied with established genome-wide thresholds, do not reliably distinguish pathogenic and benign missense variants in *PKD1*.

## Introduction

Autosomal Dominant Polycystic Kidney Disease (ADPKD) is the most common monogenic cause of kidney failure, with an estimated prevalence of 1 in 1000^1^. Typical ADPKD is largely caused by disease-causing variants in the *PKD1* and *PKD2* genes. There is genotype-phenotype correlation, in that patients with *PKD1* variants reach kidney failure on average 20 years earlier than those with *PKD2* variants^2^. Remarkably for such a common and morbid disease, the exact mechanism by which kidney cysts arise from disease-causing variants in these genes remains uncertain^3^. *PKD1* and *PKD2* encode for polycystin-1 and polycystin-2 respectively. The transmembrane regions of these two proteins form a complex for which the physiological function is not clearly defined^4^.

Approximately 85% of typical ADPKD is attributed to disease-causing variants in the *PKD1* gene^5^. The majority of disease-causing variants reported in *PKD1* are loss of function variants, though missense variants account for approximately 20%^6^. The classification of missense variants is a significant challenge in *PKD1*. Most disease-causing variants are unique to an individual family^6^ and there is at least a ten percent *de novo* rate^7^. Wide segregation analysis is not commonly possible for this adult-onset, autosomal dominant disease. A functional assay has been recently reported, but is not yet available or validated for clinical use^8^. Given the lack of other lines of evidence, *in silico* tools for prediction of pathogenicity are strongly relied upon for classification of variants in *PKD1*.

As with other monogenic diseases, the increasing burden of variants of uncertain significance is a significant challenge^9^. The need for a definitive genetic diagnosis is however increasing. Genetic diagnosis is increasingly utilised for reproductive decision making and prognostication^10,11^. Gene therapies targeted at patients with *PKD1*-disease causing variants are in clinical trial, increasing the importance of accurate identification of families with *PKD1*-mediated disease^12,13^.

There has been limited systematic study of missense variation in *PKD1*. Many *PKD1* variants reported in the literature and disease databases have been classified prior to the wide use of the American College of Medical Genetics and Genomics/Association for Molecular Pathology (ACMG/AMP) Guidelines on Variant Classification. Collating a comprehensive dataset of definitively pathogenic variants in *PKD1* would provide a valuable reference set for diagnostic and research laboratories classifying *PKD1* variants and striving to understand disease pathogenesis. This dataset would also be a valuable ‘truth’ set for validation of functional assays and for assessment of the effectiveness of heavily relied upon *in silico* tools. There is increasing evidence from assessment of other genes that genome-wide thresholds for established *in silico* tools, such as REVEL or AlphaMissense, are not always applicable to specific disease-genes, which require gene-specific calibration^14^. *PKD1* has characteristics that may challenge genome-wide rules, including strong sequence homology to a set of pseudogenes^15^, lack of a mutational hot-spot and limited knowledge of protein function.

In this study, we aimed to re-assess *PKD1* missense variants reported in variant databases and the literature and re-classify these variants using current variant classification guidelines. We then utilised this data to evaluate the efficacy of *in silico* tools in predicting pathogenicity of *PKD1* missense variants.

## Methods

All variants in the Autosomal Dominant Polycystic Kidney Disease (ADPKD) Database (PKDB) were downloaded on 16/01/2026 (https://pkdb.mayo.edu/) and filtered for *PKD1* (HGNC:9008) germline missense variants. *PKD1* germline missense variants were downloaded from ClinVar (https://www.ncbi.nlm.nih.gov/clinvar) on 23/01/2026 (transcript: NM_001009944.3) (Figure 1). *PKD1* missense variants from both databases were then input to Ensembl’s Variant Effect Predictor (VEP). GnomAD allele frequency (AF), SIFT (Sorting Intolerant From Tolerant) score, PolyPhen-2 (Polymorphism Phenotyping v2) score, CADD (Combined Annotation Dependent Depletion) score and REVEL (Rare Exome Variant Ensemble Learner) score were obtained via VEP. AlphaMissense scores across *PKD1* were downloaded from https://alphamissense.hegelab.org/. These scores were merged with both the PKDB and ClinVar variant sets.

**Figure 1.**
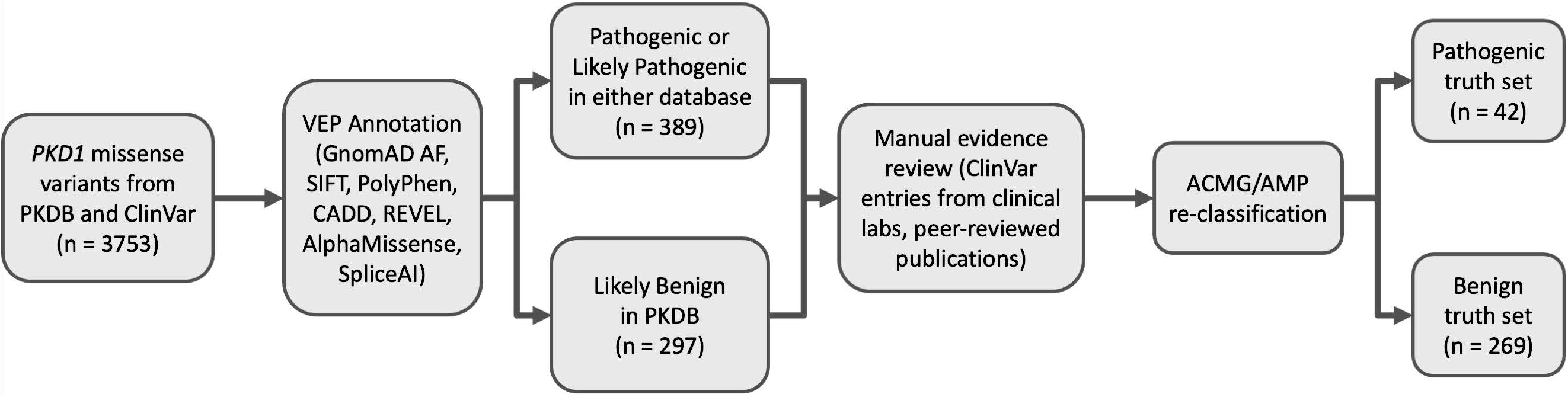
Variant extraction and curation methods workflow diagram.

The PKDB and ClinVar variants were then collated, combining duplicate variants reported across both databases, and filtered for a ‘pathogenic’ or ‘likely pathogenic’ classification in either or both databases. The source evidence for each unique variant was then systematically evaluated, including review of the original peer-reviewed publication where applicable, and a review of all ClinVar entries. Each variant was then re-classified using the ACMG/AMP Guidelines for Variant Classification^16^. The set of variants classified as ‘likely benign’ in PKDB was also extracted and re-classified using the ACMG/AMP Guidelines. The specific criteria for supporting, moderate and strong levels of evidence were determined for ADPKD based on recent literature^17–19^. The full criteria used can be found in Supplementary Table 1. SpliceAI masked scores were obtained for each of the ACMG/AMP pathogenic and likely pathogenic variants through https://spliceailookup.broadinstitute.org/, and variants predicted to alter splicing (score >0.2) were excluded from subsequent analyses.

Pathogenicity prediction performance for *PKD1* variants was assessed for the SIFT, PolyPhen-2, CADD, REVEL and AlphaMissense *in silico* tools. The distribution of pathogenicity scores was evaluated using the set of ACMG/AMP pathogenic and likely pathogenic variants (n=42), and the set of ACMG/AMP benign and likely benign variants (n=269) from the re-classification above (‘truth sets’). The thresholds used to determine pathogenic or benign supporting evidence (PP3/BP4) were derived from Pejaver et al. (2022) and Bergquist et al. (2025). For the truth set of benign variants, variants classified as likely benign in the PKDB database were chosen for evaluation rather than common (allele frequency >5%) population variants, as these variants were considered more representative of the variants that clinical testing laboratories classify with ACMG/AMP criteria and *in silico* tools.

The *PKD1* gene encodes the 4,303-amino acid polycystin-1 (PC1) protein, which has 81,757 theoretically possible missense variants. Of these, 22,879 variants arise from single-nucleotide variants and are annotated in the Ensembl database. CADD, REVEL and AlphaMissense scores were extracted for these 22,879 variants across *PKD1*. Due to limitations in the prediction tools, scores for missense variants resulting from multi-nucleotide variants were not obtained. For each *in silico* tool, the minimum, maximum and mean scores were calculated for every amino acid position.

Receiver operating characteristic (ROC) analyses were performed in R using the pROC package^20^ to evaluate the ability of CADD, REVEL, and AlphaMissense to distinguish pathogenic and benign *PKD1* missense variants. For each prediction tool, ROC curves were constructed using the same set of pathogenic and benign variants, and classification performance was summarised using the area under the curve (AUC). Confidence intervals for sensitivity across the ROC curves were estimated by bootstrap resampling using the ci.se function in pROC, while confidence intervals for AUC values were calculated using the DeLong method^21^ implemented in the ci.auc function. Pathogenic score thresholds were optimised to correspond to predefined specificities (100%, 99%, 98%, and 97%) and benign score thresholds were optimised to correspond to predefined sensitivities (100%, 99%, 98% and 97%). The true positive and false positive rates for each of the current genome-wide benign and pathogenic thresholds for each tool (derived from Pejaver et al. 2022 and Bergquist et al. 2025) were also calculated for comparison. ROC curves, confidence intervals, and threshold-specific operating points were visualized for each tool, and previously established clinical thresholds were included for reference.

To investigate circularity bias related to the use of computational evidence from REVEL (PP3 and BP4 criteria in the ACMG/AMP Guidelines) in the variant classification, the ROC analysis was repeated with a subset of variants with pathogenic or benign classification independent of PP3/BP4 evidence. This subset comprised 29 pathogenic/likely pathogenic and 208 benign/likely benign variants. For each tool, the difference in AUC between the full and subset analyses was assessed using a bootstrap permutation test (5,000 replicates) implemented in the roc.test function in pROC, with a fixed random seed set for reproducibility. As the two analyses used overlapping variant sets, the bootstrap method was used in preference to the DeLong method, which requires paired or independent observations.

## Results

A total of 3,753 missense variants in *PKD1* were obtained from PKDB and ClinVar. From this set of variants, 389 variants (10%) were reported in either or both databases as ‘Pathogenic’ or ‘Likely Pathogenic’ (Figure 2A). A total of 297 variants were reported in PKDB as ‘Likely Benign’. The original source evidence was reviewed for each of these variants, including evidence from clinical testing laboratories entered in ClinVar as well as >80 peer-reviewed publications. Eighty-nine percent (348/389) of the pathogenic variants were reported in published literature.

**Figure 2.**
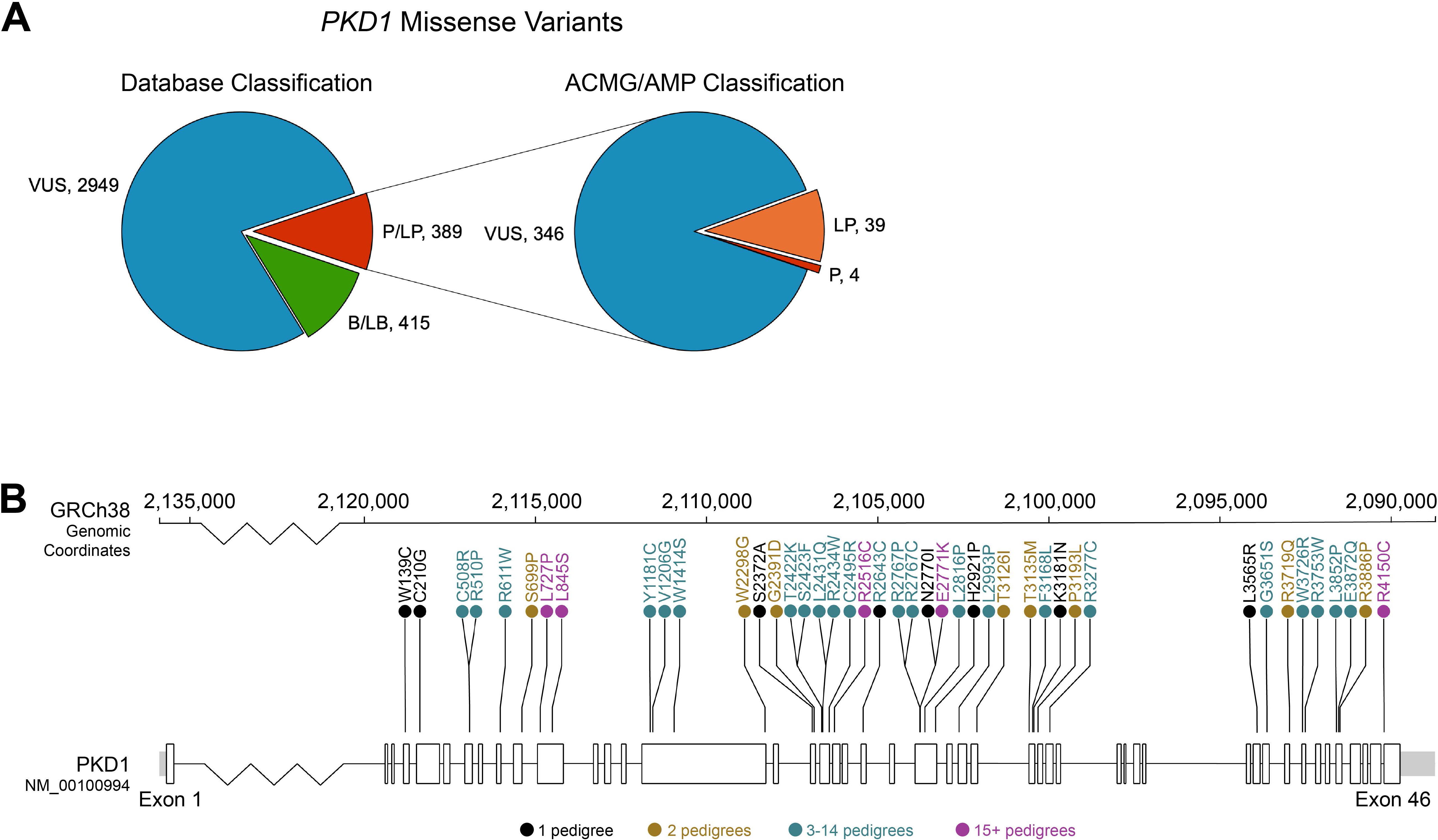
A) The number of *PKD1* missense variants by classification reported across the PKDB and ClinVar databases. Left: The red segment represents 389 variants reported as pathogenic or likely pathogenic in either database, which were collated for re-classification. Right: Re-classification of previously reported pathogenic or likely pathogenic variants following the application of the ACMG/AMP guidelines. B/LB = Benign/Likely Benign; VUS = Variant of Unknown Significance; P/LP = Pathogenic/Likely Pathogenic; LP = Likely Pathogenic; P = Pathogenic. B) Location of the 43 ACMG/AMP pathogenic or likely pathogenic missense variants along the *PKD1* gene; band at the top indicates the genomic coordinates (GRCh38). The variants in black were present in a single pedigree, the variants in gold were present in 2 pedigrees, the variants in teal were present in 3-14 pedigrees, and the variants in purple were present in 15+ pedigrees. Graphic adapted from Protein Paint by St. Jude Children’s Research Hospital.

Current ACMG/AMP Guidelines were applied to these variants (Supplementary Table 1). Of 389 reported pathogenic variants, 346 variants (89%) were downgraded to a Variant of Unknown Significance (VUS) (Figure 2A). Of the variants downgraded to VUS, 28/346 (8%) variants were reported in 3 or more unrelated pedigrees, and 52/346 (15%) had evidence from at least one segregation. Fifty-two percent (179/346) of the downgraded variants met *in silico* evidence criteria for pathogenicity based on a REVEL score >0.65^17,19^. Of 297 likely benign variants from PKDB, 269 variants (91%) met the ACMG/AMP criteria for either benign or likely benign classification and 28 variants (9%) were regraded to a VUS.

Eleven percent (43/389) of historically reported pathogenic variants met the criteria for pathogenic (four variants) or likely pathogenic (39 variants) classification using the ACMG/AMP Guidelines for Variant Interpretation (Figure 2A). Of these pathogenic and likely pathogenic variants, 28/43 (65%) are reported in 3 or more unrelated pedigrees, 33/43 (77%) had evidence from at least one segregation, and 31/43 (72%) of the variants were absent from GnomAD v4.1. Eleven of the pathogenic or likely pathogenic variants (11/43; 26%) have pathogenicity evidence from a functional study^8,22,23^. 26/43 (60%) of these pathogenic variants received PP3 *in silico* supporting evidence for pathogenicity. The 43 pathogenic missense variants were situated across most exons of the *PKD1* gene, with no mutational hotspot identified (Figure 2B). One variant (NP_001009944.3:p.(Arg3719Gln)) had SpliceAI scores greater than 0.2, and is known to alter splicing^24^, so it was subsequently excluded from further analyses due to the separate pathogenic mechanism.

To evaluate the performance of *in silico* prediction tools for *PKD1* variants, scores were obtained from SIFT^25^, PolyPhen-2^26^, CADD^27^, REVEL^28^ and AlphaMissense^29^ for the set of pathogenic/likely pathogenic *PKD1* missense variants (n=42) as well as the set of benign/likely benign *PKD1* missense variants (n=269) from the re-classification above. For the pathogenic variants, based on the current established thresholds, 62% of the variants have a REVEL score above 0.65 (n = 26) and meet the ACMG/AMP PP3 criteria for supporting evidence of pathogenicity (Figure 3A). SIFT and PolyPhen-2 had the highest number of scores meeting PP3 criteria for supporting evidence of pathogenicity (88% and 69% using thresholds of 0.999 and 0.98 respectively). CADD had 52% of pathogenic variants (n = 22) with a score above the established genome-wide threshold for pathogenicity (25.3), and AlphaMissense had 40% of pathogenic variants (n = 17) with a score above the established genome-wide threshold for pathogenicity (0.79).

**Figure 3.**
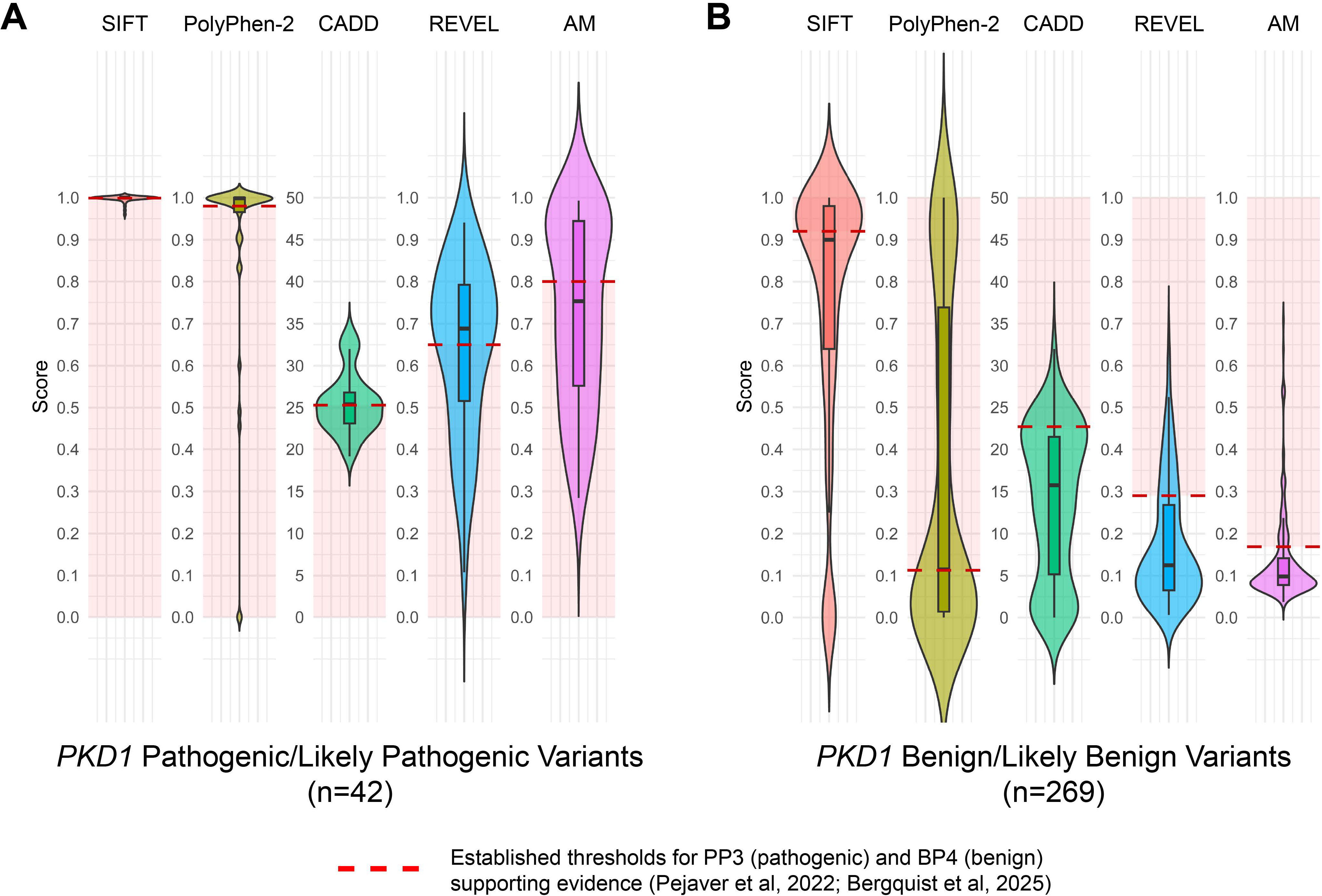
A) Distribution of scores from *in silico* prediction tools for ACMG pathogenic and likely pathogenic missense variants (n=42). The dashed red line indicates the score threshold to provide PP3 supporting evidence for ACMG pathogenic classification according to the ClinGen Working Group^17,19^. Variants falling within the red-shaded region below the defined threshold represent pathogenic variants lacking PP3 evidence to support pathogenicity. B) Distribution of scores from *in silico* prediction tools for ACMG benign and likely benign missense variants (n=269). The dashed red line indicates the score threshold to provide BP4 supporting evidence for ACMG benign classification according to the ClinGen Working Group^17,19^. Variants falling within the red-shaded region above the defined threshold represent benign variants lacking BP4 evidence to support benign classification. The SIFT scores have been inverted so that a higher score indicates a more deleterious variant. AM = AlphaMissense.

For the benign variants, based on the current established thresholds, 79% of the variants have a REVEL score below 0.29 (n = 213) and meet the ACMG/AMP BP4 criteria for supporting evidence of being benign (Figure 3B). SIFT and PolyPhen-2 had the lowest number of scores meeting BP4 criteria for supporting evidence of a benign variant (54% and 50% using thresholds of 0.92 and 0.113 respectively). Both CADD and AlphaMissense had 80% of benign variants (n = 216 and n = 215 respectively) with a score below the established genome-wide threshold for benignity (22.7 and 0.169 respectively).

To evaluate the performance of CADD, REVEL and AlphaMissense across *PKD1*, scores were obtained from each *in silico* tool for all possible single-nucleotide missense variants across *PKD1*. From 22,879 possible single-nucleotide missense variants across *PKD1* with *in silico* scores available, there was an average of five variants at each position. As seen in Figure 4, the CADD scores ranged between 0 and 34, however the scores for the set of pathogenic variants were not well distinguished from the scores for the set of benign variants. The maximum REVEL and AlphaMissense pathogenicity scores in certain regions, such as between residues 3290 and 3530 which encapsulate an unannotated segment between Transmembrane Regions 3 and 4, were consistently below the pathogenicity threshold. In the annotated functional domains, the average REVEL and AlphaMissense scores at each residue remained well below the pathogenic thresholds. Both CADD and REVEL had pathogenic variants with scores below the benign threshold as well as benign variants with scores above the pathogenic threshold. Based on these sets of variants and current genome-wide thresholds, AlphaMissense did not misclassify any of the variants.

**Figure 4.**
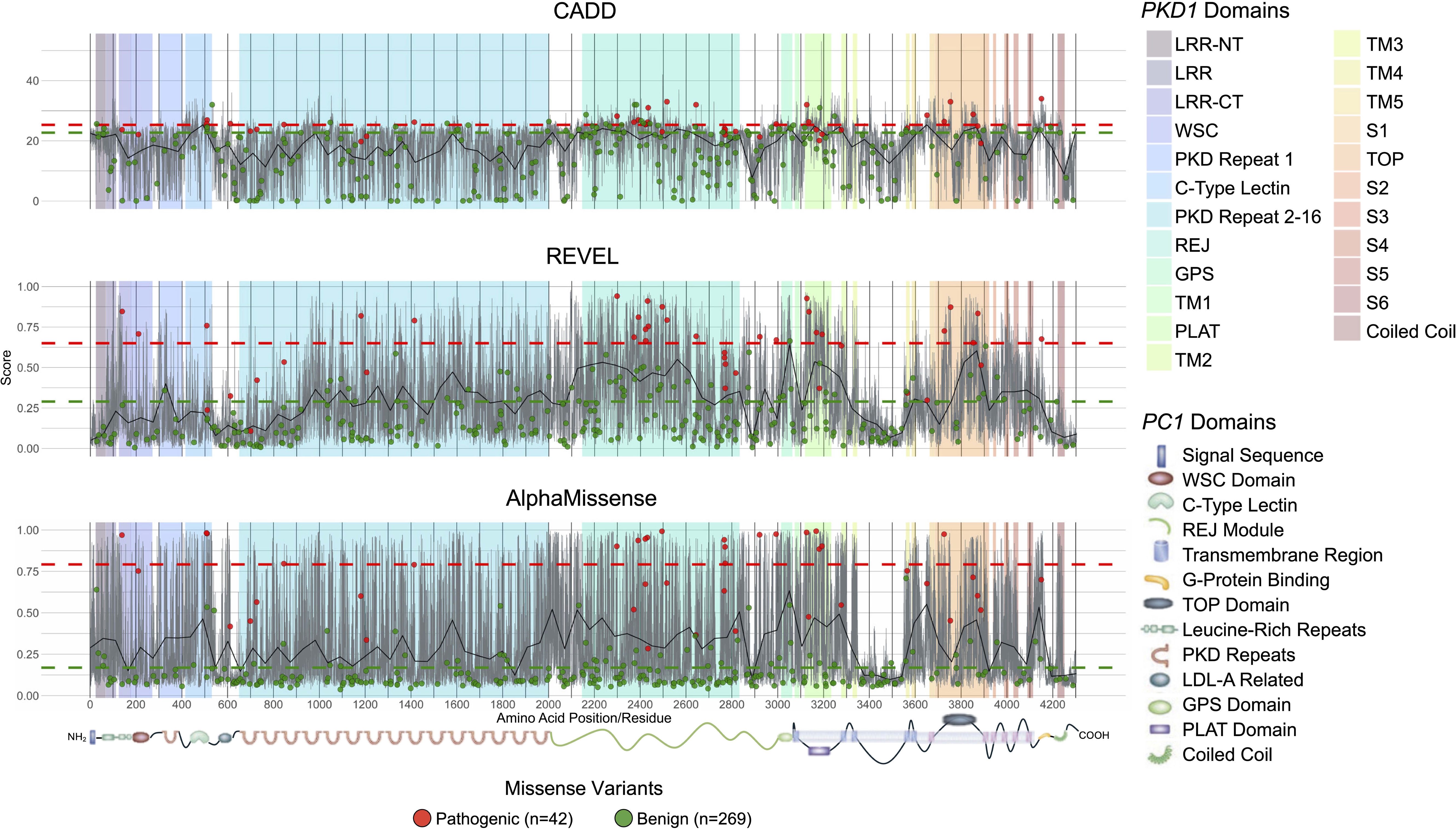
Pathogenicity scores from CADD, REVEL and AlphaMissense for single-nucleotide missense variants along *PKD1*. The functional domains are represented by coloured rectangles, and the full protein is depicted along the x axis indicating amino acid position (graphic modified from Figure 1 in Torres and Harris, 2009). The grey lines indicate the maximum and minimum scores from all possible single-nucleotide missense variants at each position. The solid black line is a smoothed average score at each position. The dashed red line is the pathogenicity threshold (CADD 25.3; REVEL 0.65; AlphaMissense 0.79) and the dashed green line is the benign threshold (CADD 22.7; REVEL 0.29; AlphaMissense 0.17) for supporting evidence for each tool from literature (Pejaver et al, 2022; Bergquist et al, 2025). The red circles represent ClinVar/PKDB variants which are pathogenic or likely pathogenic by ACMG/AMP guidelines with their relevant pathogenicity score. The green circles represent PKDB variants which are benign or likely benign by ACMG/AMP guidelines with their relevant pathogenicity score. LRR-NT = Leucine-Rich Repeats N-Terminal; LRR = Leucine-Rich Repeats; LRR-CT = Leucine-Rich Repeats C-Terminal; WSC = Wall Integrity and Stress Response Component; REJ = Receptor for Egg Jelly; GPS = G Protein-Coupled Receptor Proteolytic Site; TM = Transmembrane; PLAT = Polycystin, Lipoxygenase, and α-Toxin; S = Segment; TOP = Tetragonal Opening of Polycystins; LDL-A = Low-Density Lipoprotein A.

To determine which tool is best able to distinguish the pathogenic variants from the benign variants, a Receiver Operating Characteristic (ROC) curve analysis was performed. As shown in Figure 5, CADD had an Area Under the Curve (AUC) of 0.895 (95% Confidence Interval of 0.856-0.934), REVEL had an AUC of 0.956 (95% CI of 0.923-0.989), and AlphaMissense had the highest AUC of 0.990 (95% CI of 0.983-0.998). The sensitivity and specificity achieved by each tool using the current established clinical thresholds for PP3/BP4 supporting evidence are plotted along with the pathogenic thresholds required to achieve 100%, 99%, 98% and 97% specificity and the benign thresholds required to achieve 100%, 99%, 98% and 97% sensitivity. As the pathogenic thresholds decrease and the benign thresholds increase, the true positive rate increases but the false positive rate also increases.

**Figure 5.**
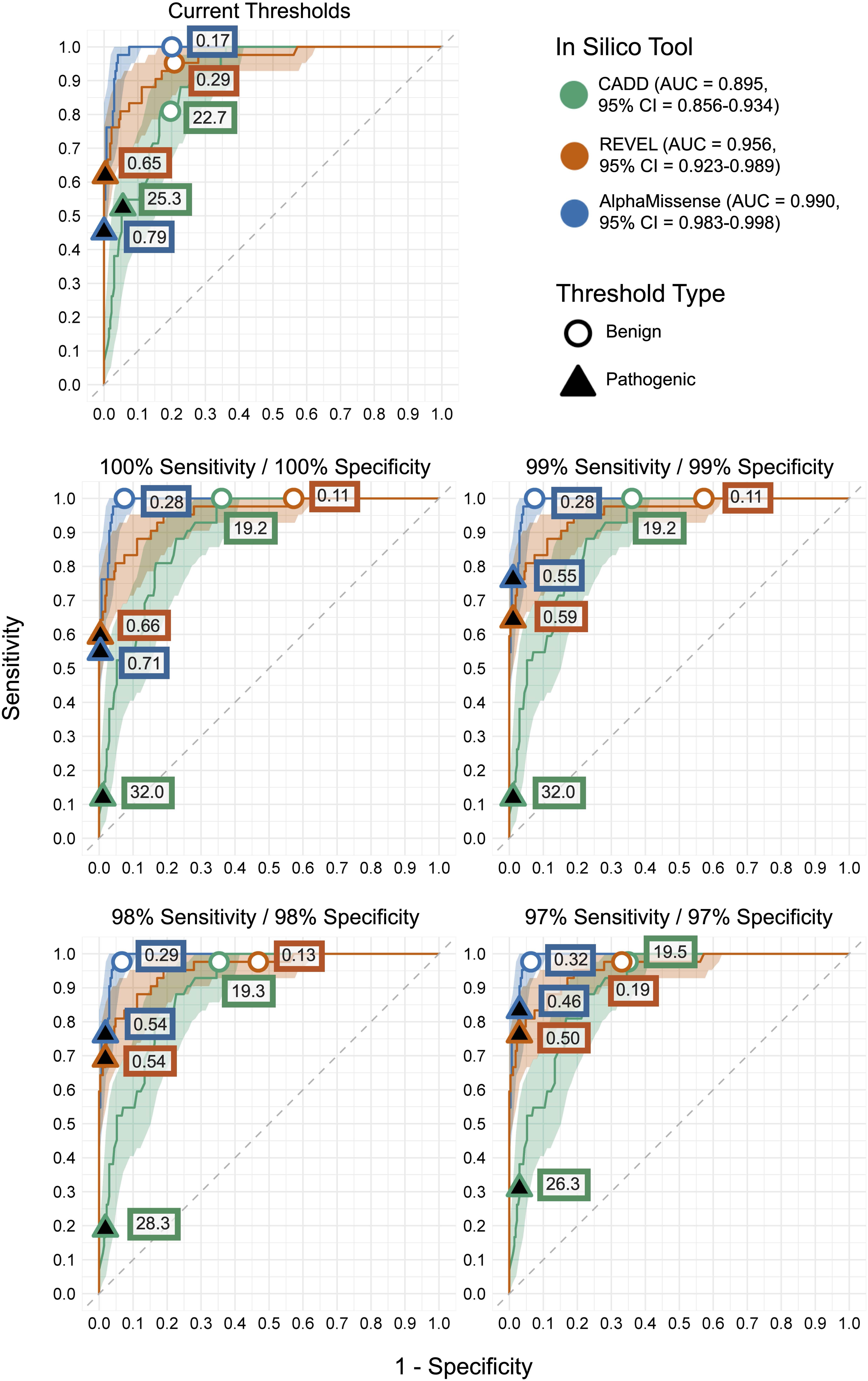
Receiver Operating Characteristic (ROC) curves for *in silico* prediction tools applied to *PKD1* pathogenic and benign missense variants. ROC curves show the relationship between sensitivity and specificity for CADD, REVEL and AlphaMissense. Shaded regions represent confidence intervals around each ROC curve. Area Under the Curve (AUC) values with 95% Confidence Intervals (CI) are specified in the legend for each tool. The diagonal dashed grey line represents random classification performance. Each panel corresponds to the current established thresholds for PP3 and BP4 criteria, as well as the pathogenic thresholds required to achieve predefined target specificity (100%, 99%, 98% and 97%) and the benign thresholds required to achieve predefined target sensitivity (100%, 99%, 98% and 97%). Labels denote the corresponding thresholds for classifying a variant as pathogenic or benign for each tool.

For the current pathogenic thresholds, REVEL (threshold 0.65) achieves the highest sensitivity of 62% true positives with a false positive rate (benign variants with scores above the pathogenic threshold) of 0.4%. If the threshold is adjusted to achieve 100% specificity (threshold of 0.66 means that no benign variants are above the pathogenic threshold), this returns 60% true positives. In comparison, the current AlphaMissense pathogenic threshold of 0.79 achieves a sensitivity of 40% true positives with 0% false positives. This threshold can be reduced to 0.71 and maintains 0% false positives, which returns 55% true positives. If modelling the pathogenic thresholds for 99%, 98% and 97% specificity, AlphaMissense achieves the highest sensitivities of 76%, 76% and 83% with thresholds of 0.55, 0.54 and 0.46 respectively.

With a benign variant achieving a score below the benign threshold considered a true negative, for the current benign thresholds, both CADD (threshold 22.7) and AlphaMissense (threshold 0.17) achieve a specificity of 80%. REVEL (threshold 0.29) achieves a specificity of 79%, so there is negligible difference in performance between the three tools. However, when modelling the benign thresholds to achieve 100% sensitivity (zero pathogenic variants below the benign threshold), the REVEL tool threshold lowers to 0.11 and returns a true negative rate of 43%. In comparison, the AlphaMissense benign threshold with zero false negatives increases to 0.28 and returns a 93% true negative rate.

The ROC curve analysis was repeated with the subset of variants achieving pathogenic/likely pathogenic (n=29) or benign/likely benign (n=208) classification with the ACMG/AMP Guidelines independent of PP3/BP4 criteria. Though the area under the curve (AUC) achieved by REVEL decreased further than the other two tools, this difference was not statistically significant (Supplementary Table 5).

## Discussion

We show that the majority of *PKD1* missense variants currently reported as disease-causing in variant databases do not meet contemporary classification guidelines for pathogenicity: 89% of historically reported pathogenic/likely pathogenic variants in PKDB and ClinVar were downgraded to Variants of Unknown Significance (VUS) using current ACMG/AMP classification criteria. We subsequently show that widely used computational pathogenicity prediction tools perform sub-optimally for *PKD1* variants when applied with established genome-wide thresholds^17,19^. Our findings highlight the ongoing difficulties in variant classification in ADPKD which is hampered by limited segregation data, a majority of disease-causing variants being private to each pedigree with no mutational hotspot, and a scarcity of functional assay data. This results in a very large majority of missense variants being classified as VUS. Molecular diagnosis of ADPKD is becoming increasingly important as the development of gene therapies progresses closer to clinical utility. Our analysis highlights the need for consistent and detailed variant reporting as well as further development of reproducible functional assays, combining genomic and functional data in order to increase pathogenic and benign classifications of missense variants in *PKD1* and improve diagnostic accuracy for ADPKD. Relying on a set of 42 ACMG/AMP pathogenic and likely pathogenic missense variants across *PKD1* is a significant limitation for all current ADPKD research, but the challenges in obtaining independent points of evidence for variant pathogenicity are ongoing.

Given the significant challenges in obtaining other lines of evidence, computational variant pathogenicity prediction tools are heavily utilised for the classification of variants in ADPKD. Using the pathogenic (n = 42) and benign (n = 269) sets of re-classified variants, we investigated the performance of widely used *in silico* tools for *PKD1* missense variants. Established genome-wide thresholds for each tool from recent literature^17,19^ were applied. It was found that SIFT and PolyPhen-2 struggled to distinguish between the pathogenic and benign variants, with a tendency to predict many of the variants as pathogenic (69-88% of confirmed pathogenic variants received a score above the pathogenic thresholds and met PP3 criteria, but 46-50% of confirmed benign variants received a score above the benign thresholds and failed BP4 criteria).

CADD scored the benign variants more accurately (80% specificity), but still overall struggled to distinguish benign and pathogenic variants. When *in silico* scores for all possible single-nucleotide missense variants across the PC1 protein were plotted by residue (Figure 4), CADD did not clearly separate the pathogenic and benign variants at any position. The tool produced bidirectional misclassifications, with pathogenic variants below the benign threshold and benign variants above the pathogenic threshold.

REVEL and AlphaMissense both achieved considerably higher discrimination between the pathogenic and benign variants, with areas under the curve (AUC) greater than 95% in the ROC curve analysis (Figure 5). Based on current genome-wide thresholds, REVEL achieved the highest sensitivity of 62% with a low false positive (benign variants with a score above the pathogenic threshold) rate of 0.4%, however a significant number of pathogenic variants fail PP3 criteria. Notably, REVEL also output a few variant misclassifications (2 pathogenic variants and 1 benign variant). AlphaMissense didn’t misclassify any of the variants included in this analysis (i.e. score an established pathogenic variant as benign or vice versa). However, AlphaMissense has lower sensitivity and specificity due to a large number of variants (25%) with an ambiguous score between the pathogenic and benign thresholds. Based on all missense variants across *PKD1* (Figure 4), the average REVEL and AlphaMissense scores, even across annotated functional domains, remained well below the pathogenic thresholds, which may explain the low sensitivities observed. These results have shown that applying the current genome-wide thresholds for these tools to *PKD1* missense variants results in substantial loss of either sensitivity or specificity.

The variable performance of these *in silico* tools for missense variants across *PKD1* using genome-wide thresholds is likely due to a combination of factors. Firstly, *PKD1* is likely underrepresented in the training data for these tools. REVEL was trained using HGMD variants reported between 2012-15^28^, a period when *PKD1* genetic testing was technically challenging and disease-specific databases such as PKDB were the primary repository for reported variants rather than general databases such as HGMD^30^.

Consistent with our findings, clinical PKD testing laboratories have independently flagged REVEL performance issues in *PKD1*^31^. AlphaMissense is not reliant on previously classified pathogenic and benign variants like REVEL, however is reliant on accurate protein structure prediction and population variant frequency data^29^. The entire structure of the large 4,303 amino acid PC1 protein is unsolved^4^, and the predicted structure from the AlphaFold Server has a predicted template modelling (pTM) score of 0.42, which indicates the predicted structure is likely to be incorrect^32^. Population variant frequency data for *PKD1* likely underrepresents variants due to challenges of mapping short-read sequencing data across a large portion of *PKD1* that has very high (>97%) sequence homology to a set of pseudogenes^33^. As such, *PKD1* is likely not well represented in the training datasets for any of the *in silico* tools, which greatly reduces the accuracy and reliability of the output scores.

*PKD1* is also likely underrepresented in the dataset used to calibrate the genome-wide thresholds for these computational tools. The ClinGen Sequence Variant Interpretation Working Group utilised a probabilistic model to generate genome-wide cut-offs for strengths of evidence (ranging from very strong to supporting) for a range of computational variant prediction tools. The variant data used for this genome-wide calibration excluded variants in segmental duplications^17^. The homologous *PKD1* pseudogenes are a result of a segmental duplication event in primate evolution^34^. Therefore, it is likely that most *PKD1* variants would have been excluded from this genome-wide calibration. This reduced calibration data may explain some of the discordance in evidence strengths attributed to *PKD1* variants, as exemplified by some benign *PKD1* missense variants in our dataset receiving CADD scores above the Pejaver et al (2022) threshold for moderate level evidence of pathogenicity. Previous studies have already established that these *in silico* tools can behave differently in different genes depending on their size, structure and function^35,36^ and that genome-wide calibrated thresholds should not be used for genes where the distribution of predicted scores is skewed^14,37^. The lack of *PKD1* relevant data used in establishing these *in silico* tools and thresholds for their application suggests that the established genome-wide thresholds for these tools may not be applicable to *PKD1* missense variants.

Given the *PKD1*-specific challenges of accurate variant classification, we explored adjusting the tool thresholds to improve the performance of *in silico* tools for *PKD1* missense variant classification. There is precedent for calculating gene-specific thresholds for *in silico* tools in other diseases, such as *PAXc* and *BRCA1*^35,38,39^. The high AUCs achieved by REVEL and AlphaMissense in our ROC curve analysis indicate that both tools have the capacity to discriminate between pathogenic and benign *PKD1* variants, but optimal thresholds need to be calculated to do so with high sensitivity and specificity. However, using our definitive sets of pathogenic and benign *PKD1* variants, no threshold adjustment that was modelled (Figure 5) was able to achieve high sensitivity with a clinically acceptable false-positive rate. Our analysis was limited by the small size of our truth set of definitive pathogenic and benign variants. There is also potential bias in our dataset since REVEL was used as the computational tool to classify the variants in our dataset. Therefore, the ROC curve analysis was repeated with a subset of variants receiving pathogenic or benign ACMG/AMP classification independent of PP3/BP4 evidence and the difference in the AUC for all three tools was not significant (Supplementary Table 5), acknowledging this was a smaller dataset. The current truth set of variants are likely too small for calculating a gene-specific threshold calibration that can suitably balance sensitivity and specificity for clinical use. However, with sufficient and balanced data in the future, a gene-specific threshold calibration would likely improve the utility of *in silico* tools for pathogenicity predictions of missense variants in *PKD1*.

Accurate classification of *PKD1* missense variants is becoming increasingly important, as molecular diagnosis informs prognosis, reproductive planning, and eligibility for clinical trials. Based on current ACMG/AMP Guidelines, a large majority of missense variants in *PKD1* are classified as VUS, with only 43 variants across the gene receiving pathogenic or likely pathogenic classification. This is a remarkably low number when it is considered that ADPKD has a prevalence of 1/1000^1^, that most variants are private to each pedigree, and ∼20% of variants are missense variants^6^. It is unlikely that the upcoming ACMG/AMP V4 variant classification guidelines will significantly alter the proportion of VUS in *PKD1*, as we have already applied contemporary adjustments to the original guidelines. If the newest update increases weighting of *in silico* tool evidence, this may be particularly concerning for *PKD1*, where our data suggests caution with reliance on *in silico* scores. Our data has highlighted the need for accurate collation of pedigree segregation, cohort and clinical laboratory data as well as consistent and detailed reporting of variants to expand the ACMG/AMP pathogenic and benign variant sets in *PKD1*. Equally, it highlights the value of continued discovery science efforts to better understand the function of poycystin-1 and relatedly develop high-throughput functional assays^8^ to provide PS3/BS3 evidence for missense variants. This combination of genomic and functional data will enable future gene-specific threshold calibration of *in silico* tools with a more comprehensive dataset and will be key to improving diagnostic accuracy in ADPKD.

## Code Availability

The code used in this analysis is available on GitHub at https://github.com/nlehmann-beep/PKD1_missense_variant_analysis and archived on Zenodo at https://doi.org/10.5281/zenodo.21637865.

## Author Contributions

Conceptualization: A.M., G.R.; Data Curation: S.K., N.L., Y.H.; Formal analysis: N.L., S.K.; Funding acquisition: A.M.; Methodology: A.M., R.R, G.H.; Supervision: A.M., Writing-original draft N.L., A.M.; Writing-review C editing: A.M., S.K., G.R., R.R., G.H.

## Data Availability

All data produced in the present study are contained in the supplementary tables, and the code used in the analysis is available on Zenodo at https://doi.org/10.5281/zenodo.21637865.

## References

1. Lanktree MB, Haghighi A, Guiard E, et al. Prevalence Estimates of Polycystic Kidney and Liver Disease by Population Sequencing. J Am Soc Nephrol. 2018;29(10):2593–2600. doi:10.1681/ASN.2018050493

2. Cornec-Le Gall E, Audrézet MP, Chen JM, et al. Type of PKD1 Mutation Influences Renal Outcome in ADPKD. J Am Soc Nephrol. 2013;24(6):1006–1013. doi:10.1681/ASN.2012070650

3. Mallawaarachchi AC, Hort Y, Wedd L, et al. Somatic mutation in autosomal dominant polycystic kidney disease revealed by deep sequencing human kidney cysts. npj Genom Med. 2024;9(1):69. doi:10.1038/s41525-024-00452-6

4. Su Q, Hu F, Ge X, et al. Structure of the human PKD1-PKD2 complex. Science. 2018;361(6406):eaat9819. doi:10.1126/science.aat9819

5. Torres VE, Harris PC, Pirson Y. Autosomal dominant polycystic kidney disease. The Lancet. 2007;369(9569):1287–1301. doi:10.1016/S0140-6736(07)60601-1

6. Audrézet MP, Cornec-Le Gall E, Chen JM, et al. Autosomal dominant polycystic kidney disease: Comprehensive mutation analysis of PKD1 and PKD2 in 700 unrelated patients. Human Mutation. 2012;33(8):1239–1250. doi:10.1002/humu.22103

7. Neumann HPH, Jilg C, Bacher J, et al. Epidemiology of autosomal-dominant polycystic kidney disease: an in-depth clinical study for south-western Germany. Nephrol Dial Transplant. 2013;28(6):1472–1487. doi:10.1093/ndt/gfs551

8. Ha K, Loeb GB, Park M, et al. Disruption of Polycystin Ciliary Localization and Channel Function by Autosomal Dominant Polycystic Kidney Disease-Causing Polycystin-1 Variants. J Am Soc Nephrol. Published online February 10, 2026. doi:10.1681/ASN.0000001008

9. Yang H, Sieben CJ, Schauer RS, Harris PC. Genetic Spectrum of Polycystic Kidney and Liver Diseases and the Resulting Phenotypes. Adv Kidney Dis Health. 2023;30(5):397–406. doi:10.1053/j.akdh.2023.04.004

10. Devuyst O, Ahn C, Barten TRM, et al. KDIGO 2025 Clinical Practice Guideline for the Evaluation, Management, and Treatment of Autosomal Dominant Polycystic Kidney Disease (ADPKD). Kidney International. 2025;107(2):S1–S239. doi:10.1016/j.kint.2024.07.009

11. Soraru J, Mallett AJ, McCarthy H, Mallawaarachchi A. Genetic Testing in Cystic Kidney Disease. Kidney360. Published online January 2, 2026. doi:10.34067/KID.0000001127

12. Lakhia R, Alvarez J, Ramalingam H, et al. RGLS8429-Mediated miR-17 Inhibition Leads to Acute PKD1/2 De-repression and Ameliorates Preclinical ADPKD: TH-PO435. Journal of the American Society of Nephrology. 2024;35. doi:10.1681/ASN.2024fvfc3k6z

13. Stirnweiss A, Chakera A, Stevenson J, et al. WCN26-6082 A PHASE 1 CLINICAL TRIAL OF PYC-003 FOR THE TREATMENT OF PATIENTS WITH AUTOSOMAL DOMINANT POLYCYSTIC KIDNEY DISEASE (ADPKD). Kidney International Reports. 2026;11(4). doi:10.1016/j.ekir.2026.106401

14. Tejura M, Fayer S, McEwen AE, Flynn J, Starita LM, Fowler DM. Calibration of variant effect predictors on genome-wide data masks heterogeneous performance across genes. Am J Hum Genet. 2024;111(9):2031–2043. doi:10.1016/j.ajhg.2024.07.018

15. Mallawaarachchi AC, Lundie B, Hort Y, et al. Genomic diagnostics in polycystic kidney disease: an assessment of real-world use of whole-genome sequencing. Eur J Hum Genet. 2021;29(5):760–770. doi:10.1038/s41431-020-00796-4

16. Richards S, Aziz N, Bale S, et al. Standards and guidelines for the interpretation of sequence variants: a joint consensus recommendation of the American College of Medical Genetics and Genomics and the Association for Molecular Pathology. Genet Med. 2015;17(5):405–424. doi:10.1038/gim.2015.30

17. Pejaver V, Byrne AB, Feng BJ, et al. Calibration of computational tools for missense variant pathogenicity classification and ClinGen recommendations for PP3/BP4 criteria. Am J Hum Genet. 2022;109(12):2163–2177. doi:10.1016/j.ajhg.2022.10.013

18. Biesecker LG, Byrne AB, Harrison SM, et al. ClinGen guidance for use of the PP1/BS4 co-segregation and PP4 phenotype specificity criteria for sequence variant pathogenicity classification. Am J Hum Genet. 2024;111(1):24–38. doi:10.1016/j.ajhg.2023.11.009

19. Bergquist T, Stenton SL, Nadeau EAW, et al. Calibration of additional computational tools expands ClinGen recommendation options for variant classification with PP3/BP4 criteria. Genetics in Medicine. 2025;27(6):101402. doi:10.1016/j.gim.2025.101402

20. Robin X, Turck N, Hainard A, et al. pROC: an open-source package for R and S+ to analyze and compare ROC curves. BMC Bioinformatics. 2011;12:77. doi:10.1186/1471-2105-12-77

21 DeLong ER, DeLong DM, Clarke-Pearson DL. Comparing the Areas under Two or More Correlated Receiver Operating Characteristic Curves: A Nonparametric Approach. Biometrics. 1988;44(3):837–845. doi:10.2307/2531595

22. Qian F, Boletta A, Bhunia AK, et al. Cleavage of polycystin-1 requires the receptor for egg jelly domain and is disrupted by human autosomal-dominant polycystic kidney disease 1-associated mutations. Proceedings of the National Academy of Sciences. 2002;99(26):16981–16986. doi:10.1073/pnas.252484899

23. Cai Y, Fedeles SV, Dong K, et al. Altered trafficking and stability of polycystins underlie polycystic kidney disease. J Clin Invest. 2014;124(12):5129–5144. doi:10.1172/JCI67273

24. Gonzalez-Paredes FJ, Ramos-Trujillo E, Claverie-Martin F. Defective pre-mRNA splicing in *PKD1* due to presumed missense and synonymous mutations causing autosomal dominant polycystic disease. Gene. 2014;546(2):243–249. doi:10.1016/j.gene.2014.06.004

25. Sim NL, Kumar P, Hu J, Henikoff S, Schneider G, Ng PC. SIFT web server: predicting effects of amino acid substitutions on proteins. Nucleic Acids Res. 2012;40(Web Server issue):W452–W457. doi:10.1093/nar/gks539

26. Adzhubei I, Jordan DM, Sunyaev SR. Predicting Functional Effect of Human Missense Mutations Using PolyPhen-2. Current Protocols in Human Genetics. 2013;76(1):7.20.1–7.20.41. doi:10.1002/0471142905.hg0720s76

27. Rentzsch P, Witten D, Cooper GM, Shendure J, Kircher M. CADD: predicting the deleteriousness of variants throughout the human genome. Nucleic Acids Res. 2019;47(D1):D886–D894. doi:10.1093/nar/gky1016

28. Ioannidis NM, Rothstein JH, Pejaver V, et al. REVEL: An Ensemble Method for Predicting the Pathogenicity of Rare Missense Variants. Am J Hum Genet. 2016;99(4):877–885. doi:10.1016/j.ajhg.2016.08.016

29. Cheng J, Novati G, Pan J, et al. Accurate proteome-wide missense variant effect prediction with AlphaMissense. Science. 2023;381(6664):eadg7492. doi:10.1126/science.adg7492

30. Obeidova L, Elisakova V, Stekrova J, et al. Novel mutations of PKD genes in the Czech population with autosomal dominant polycystic kidney disease. BMC Med Genet. 2014;15:41. doi:10.1186/1471-2350-15-41

31. Durkie M, Cassidy EJ, Berry I, et al. ACGS Best Practice Guidelines for Variant Classification in Rare Disease 2024. Published online 2024.

32. Xu J, Zhang Y. How significant is a protein structure similarity with TM-score = 0.5? Bioinformatics. 2010;26(7):889–895. doi:10.1093/bioinformatics/btq066

33. Mallawaarachchi AC, Furlong TJ, Shine J, Harris PC, Cowley MJ. Population data improves variant interpretation in autosomal dominant polycystic kidney disease. Genet Med. 2019;21(6):1425–1434. doi:10.1038/s41436-018-0324-x

34. Bogdanova N, Markoff A, Gerke V, McCluskey M, Horst J, Dworniczak B. Homologues to the first gene for autosomal dominant polycystic kidney disease are pseudogenes. Genomics. 2001;74(3):333–341. doi:10.1006/geno.2001.6568

35. Tian Y, Pesaran T, Chamberlin A, et al. REVEL and BayesDel outperform other in silico meta-predictors for clinical variant classification. Sci Rep. 2019;9(1):12752. doi:10.1038/s41598-019-49224-8

36. Tordai H, Torres O, Csepi M, Padányi R, Lukács GL, Hegedűs T. Analysis of AlphaMissense data in different protein groups and structural context. Sci Data. 2024;11(1):495. doi:10.1038/s41597-024-03327-8

37. Chen Y, Fayer S, Jain S, et al. Gene- and domain-aware calibration increases the clinical utility of variant effect predictors. bioRxiv. Preprint posted online February 18, 2026:2026.02.17.706269. doi:10.64898/2026.02.17.706269

38. Andhika NS, Biswas S, Hardcastle C, et al. Using computational approaches to enhance the interpretation of missense variants in the PAX6 gene. Eur J Hum Genet. 2024;32(8):1005–1013. doi:10.1038/s41431-024-01638-3

39. Montanucci L, Brünger T, Boßelmann CM, et al. Evaluating novel in silico tools for accurate pathogenicity classification in epilepsy-associated genetic missense variants. Epilepsia. 2024;65(12):3655–3663. doi:10.1111/epi.18155

40. Jarvik GP, Browning BL. Consideration of Cosegregation in the Pathogenicity Classification of Genomic Variants. The American Journal of Human Genetics. 2016;98(6):1077–1081. doi:10.1016/j.ajhg.2016.04.003

