## Supplementary Data and Tables for "Challenges in Classification of *PKD1* Missense Variation in Autosomal Dominant Polycystic Kidney Disease"

| **Evidence Type** | **Categories** | **Applicability for**  **ADPKD Missense Variants** |
| --- | --- | --- |
| **Phenotypic** | **PP4** – Patient’s phenotype or family history is highly specific for a disease with a single genetic aetiology | **Applicable** – *PKD1* established as cause of disease, although other genes may also cause ADPKD in a minority of cases |
| **Functional Evidence** | **PS3 –** Well-established *in vitro* or *in vivo* functional studies supportive of a damaging effect on the gene or gene product | **Applicable –** downgraded to moderate level evidence, functional assays for ADPKD are unvalidated^8^ |
|  | **BS3 –** Well-established *in vitro* or *in vivo* functional studies show no damaging effect on protein function | **Applicable –** downgraded to moderate level evidence, functional assays for ADPKD are unvalidated^8^ |
| ***In-Silico*** | **PP3** – Multiple lines of computational evidence support a deleterious effect on the gene or gene product | **Applicable** – REVEL score above 0.65 used as pathogenic threshold^17,19^ |
|  | **BP4** – Multiple lines of computational evidence suggest no impact on the gene or gene product | **Applicable** – REVEL score below 0.29 used as benign threshold^17,19^ |
| **Family Studies** | **PS2** – *De novo* (both maternity and paternity confirmed) in a patient with the disease and no family history | **Applicable** |
|  | **PM3 –** For recessive disorders, detected in trans with a pathogenic variant | **Not applicable** – disease follows autosomal dominant inheritance pattern |
|  | **PM6** – Assumed *de novo*, but without confirmation of paternity and maternity | **Applicable** |
|  | **PP1** – Co-segregation with disease in multiple affected family members in a gene definitively known to cause the disease | **Applicable** – updated to supporting level for 1-3 segregations, moderate level for 4-5 segregations, and strong level for 6+ segregations^40^ |
|  | **BS4** – Non-segregation with disease | **Applicable** |
|  | **BP2** – Observed in trans (on different alleles) with a pathogenic variant for a fully penetrant dominant gene/disorder; or observed in cis (same allele) with a pathogenic variant in any inheritance pattern | **Applicable** |
| **Previous Reports** | **PS1** – Same amino acid change as a previously established pathogenic variant regardless of nucleotide change | **Applicable** |
|  | **PS4 –** The prevalence of the variant in affected individuals is significantly increased compared with the prevalence in controls | **Applicable –** updated to supporting level for 2 independent pedigrees, moderate level for 3-14 pedigrees, and strong level for 15+ pedigrees^16,31^ |
|  | **PM2 –** Absent from controls in Exome Sequencing Project, 1000 Genomes Project, or Exome Aggregation Consortium | **Applicable** – gnomAD used as proxy for controls, downgraded to supporting level evidence^31^ |
|  | **PM5 –** Novel missense change at an amino acid residue where a different missense change determined to be pathogenic has been seen before | **Applicable** |
|  | **PP2** – Missense variant in a gene that has a low rate of benign missense variation and in which missense variants are a common mechanism of disease | **Not applicable** – unknown mechanism of disease |
|  | **PP5** – Reputable source recently reports variant as pathogenic, but the evidence is not available to the laboratory to perform an independent evaluation | **Not applicable** – highly interpretive |
|  | **BA1** – Allele frequency is above 5% | **Applicable** – Allele frequency in GnomAD above 0.05 |
|  | **BS1 –** Allele frequency is greater than expected for disorder | **Applicable** – Allele Frequency (across all genomes or any subpopulation group) above 0.00013 or found in >99 alleles in GnomAD, downgrade to supporting if found in 12-99 alleles in GnomAD |
|  | **BS2 –** Observed in a healthy adult individual for a recessive (homozygous), dominant (heterozygous), or  X-linked (hemizygous) disorder with full penetrance expected at an early age | **Applicable** – Heterozygous variant observed in 2+ healthy individuals |
| **Molecular** | **PVS1 –** null variant in a gene where loss of function (LOF) is a known mechanism of disease | **Not applicable** – only missense variants reviewed |
|  | **PM1** – Located in a mutational hot spot and/or critical and well-established functional domain without benign variation | **Not applicable** – no known mutational hot spot or established functional domain in ADPKD |
|  | **PM4** – Protein length changes as a result of in-frame deletions/insertions in a nonrepeat region or stop-loss variants | **Not applicable** – only missense variants reviewed |
|  | **BP1** – Missense variant in a gene for which primarily truncating variants are known to cause disease | **Not applicable –** non-truncating variants have been shown to be pathogenic |
|  | **BP3** – In-frame deletions/insertions in a repetitive region without a known function | **Not applicable** – only missense variants reviewed |
|  | **BP5** – Variant found in a case with an alternate molecular basis for disease | **Applicable** |
|  | **BP7** – A synonymous (silent) variant for which splicing prediction algorithms predict no impact to the splice consensus sequence nor the creation of a new splice site AND the nucleotide is not highly conserved; splice variants where RNA studies have confirmed no impact | **Not applicable** – only missense variants reviewed |

*Supplementary Table 1. ACMG/AMP variant classification criteria categories, detailing whether they apply to ADPKD and any changes made to the evidence levels. The categories with a white background provide evidence for pathogenicity, and the categories with a grey background provide evidence for a benign variant classification.*

| **Exon** | **cDNA Change^a^** | **Amino Acid Change^b^** | **Database** | **Reported Classification in PKDB** | **Reported Classification**  **in ClinVar** | **ACMG Criteria Applied** | **ACMG Classification** |
| --- | --- | --- | --- | --- | --- | --- | --- |
| 4/46 | c.417G>T | p.(Trp139Cys) | PKDB | Likely Pathogenic | - | PM2-Sup,PP1-Str,PP3,PP4, PS3-Mod | LP |
| 5/46 | c.628T>G | p.(Cys210Gly) | PKDB | Likely Pathogenic | - | PS3-Mod,PM2-Sup,PP1-Sup,PP3,PP4 | LP |
| 7/46 | c.1522T>C | p.(Cys508Arg) | Both | Likely Pathogenic | Conflicting classifications of pathogenicity | PS4-Mod,PM2-Sup,PP1-Sup,PP3,PP4 | LP |
| 7/46 | c.1529G>C | p.(Arg510Pro) | ClinVar | - | Likely pathogenic | PS4-Mod,PM2-Sup,PM6,PP4 | LP |
| 9/46 | c.1831C>T | p.(Arg611Trp) | Both | Likely Pathogenic | Conflicting classifications of pathogenicity | PS4-Mod,PM2-Sup,PM6,PP4 | LP |
| 10/46 | c.2095T>C | p.(Ser699Pro) | ClinVar | - | Likely pathogenic | PS2,PS4-Sup,PM2-Sup,PP4 | LP |
| 11/46 | c.2180T>C | p.(Leu727Pro) | Both | Likely Pathogenic | Pathogenic/Likely pathogenic | PS4-Str,PM2-Sup,PM6,PP1-Sup,PP4 | LP |
| 11/46 | c.2534T>C | p.(Leu845Ser) | Both | Likely Pathogenic | Pathogenic/Likely pathogenic | PS4-Str,PP1-Str,PP4 | P |
| 15/46 | c.3542A>G | p.(Tyr1181Cys) | ClinVar | - | Likely pathogenic | PS4-Mod,PM2-Sup,PP1-Sup,PP3,PP4 | LP |
| 15/46 | c.3617T>G | p.(Val1206Gly) | PKDB | Likely Pathogenic | - | PS4-Mod,PM2-Sup,PM6,PP1-Sup,PP4 | LP |
| 15/46 | c.4241G>C | p.(Trp1414Ser) | ClinVar | - | Likely pathogenic | PS4-Mod,PM2-Sup,PP1-Sup,PP3,PP4 | LP |
| 15/46 | c.6892T>G | p.(Trp2298Gly) | Both | Likely Pathogenic | Likely pathogenic | PS4-Sup,PM2-Sup,PM6,PP1-Sup,PP3,PP4 | LP |
| 17/46 | c.7114T>G | p.(Ser2372Ala) | ClinVar | - | Likely pathogenic | PS2,PM2-Sup,PP3,PP4 | LP |
| 17/46 | c.7172G>A | p.(Gly2391Asp) | PKDB | Likely Pathogenic | - | PS4-Sup,PM2-Sup,PM6,PP3,PP4 | LP |
| 18/46 | c.7265C>A | p.(Thr2422Lys) | Both | Likely Pathogenic | Uncertain significance | PS4-Mod,PM2-Sup,PP1-Sup,PP3,PP4 | LP |
| 18/46 | c.7268C>T | p.(Ser2423Phe) | Both | Likely Pathogenic | Likely pathogenic | PS4-Mod,PM2-Sup,PP1-Sup,PP3,PP4 | LP |
| 18/46 | c.7292T>A | p.(Leu2431Gln) | ClinVar | - | Pathogenic/Likely pathogenic | PS4-Mod,PM2-Sup,PP1-Mod,PP3,PP4 | LP |
| 18/46 | c.7300C>T | p.(Arg2434Trp) | Both | Likely Pathogenic | Conflicting classifications of pathogenicity | PS4-Mod,PM2-Sup,PP1-Sup,PP3,PP4 | LP |
| 18/46 | c.7483T>C | p.(Cys2495Arg) | Both | Likely Pathogenic | Pathogenic/Likely pathogenic | PS4-Mod,PM2-Sup,PM6,PP1-Sup,PP3,PP4 | LP |
| 19/46 | c.7546C>T | p.(Arg2516Cys) | Both | Likely Pathogenic | Pathogenic/Likely pathogenic | PS4-Str,PM2-Sup,PM6,PP1-Sup,PP3,PP4 | P |
| 21/46 | c.7927C>T | p.(Arg2643Cys) | Both | Likely Pathogenic | Pathogenic/Likely pathogenic | PS4-Mod,PM6,PP3,PP4 | LP |
| 23/46 | c.8299C>T | p.(Arg2767Cys) | Both | Likely Pathogenic | Pathogenic/Likely pathogenic | PS4-Mod,PM2-Sup,PM6,PP1-Sup,PP4 | LP |
| 23/46 | c.8300G>C | p.(Arg2767Pro) | Both | Likely Pathogenic | Likely pathogenic | PS4-Mod,PM2-Sup,PP1-Str,PP4 | LP |
| 23/46 | c.8309A>T | p.(Asn2770Ile) | PKDB | Likely Pathogenic | - | PM2-Sup,PP1-Str,PP4 | LP |
| 23/46 | c.8311G>A | p.(Glu2771Lys) | Both | Likely Pathogenic | Pathogenic/Likely pathogenic | PS3-Mod,PS4-Str,PM6,PP1-Str,PP4 | P |
| 23/46 | c.8447T>C | p.(Leu2816Pro) | Both | Likely Pathogenic | Pathogenic/Likely pathogenic | PS3-Mod,PS4-Mod,PM2-Sup,PP1-Mod,PP4 | LP |
| 23/46 | c.8762A>C | p.(His2921Pro) | PKDB | Likely Pathogenic | - | PS2,PM2-Sup,PP1-Mod,PP3,PP4 | LP |
| 25/46 | c.8978T>C | p.(Leu2993Pro) | Both | Likely Pathogenic | Likely pathogenic | PS3-Mod,PS4-Mod,PM2-Sup,PM6,PP1-Sup,PP3,PP4 | LP |
| 26/46 | c.9377C>T | p.(Thr3126Ile) | Both | Likely Pathogenic | Conflicting classifications of pathogenicity | PS4-Sup,PM2-Sup,PM6,PP1-Sup,PP3,PP4 | LP |
| 27/46 | c.9404C>T | p.(Thr3135Met) | Both | Likely Pathogenic | Conflicting classifications of pathogenicity | PS4-Sup,PM2-Sup,PP3,PP4, PS3-Mod | LP |
| 27/46 | c.9504C>G | p.(Phe3168Leu) | Both | Likely Pathogenic | Conflicting classifications of pathogenicity | PS4-Mod,PM2-Sup,PP1-Sup,PP3,PP4 | LP |
| 27/46 | c.9543G>T | p.(Lys3181Asn) | ClinVar | - | Likely pathogenic | PS2,PM2-Sup,PP4 | LP |
| 28/46 | c.9578C>T | p.(Pro3193Leu) | Both | Likely Pathogenic | Uncertain significance | PS4-Sup,PM2-Sup,PP1-Mod,PP3,PP4 | LP |
| 29/46 | c.9829C>T | p.(Arg3277Cys) | Both | VUS | Pathogenic/Likely pathogenic | PS3-Mod,PS4-Mod,PP1-Str,PP4 | LP |
| 37/46 | c.10951G>A | p.(Gly3651Ser) | Both | Likely Pathogenic | Conflicting classifications of pathogenicity | PS4-Mod,PM2-Sup,PM6,PP1-Sup,PP4, PS3-Mod | LP |
| 38/46 | c.11156G>A | p.(Arg3719Gln) | Both | Likely Pathogenic | Pathogenic | PS4-Mod,PM2-Sup,PP1-Sup,PP4, PS3-Mod | LP |
| 39/46 | c.11176T>C | p.(Trp3726Arg) | Both | Likely Pathogenic | Pathogenic/Likely pathogenic | PS4-Mod,PM2-Sup,PM6,PP3,PP4 | LP |
| 39/46 | c.11257C>T | p.(Arg3753Trp) | Both | Likely Pathogenic | Pathogenic | PS4-Mod,PM2-Sup,PM6,PP1-Sup,PP3,PP4 | LP |
| 42/46 | c.11555T>C | p.(Leu3852Pro) | Both | Likely Pathogenic | Likely pathogenic | PS4-Mod,PM2-Sup,PP1-Sup,PP3,PP4 | LP |
| 42/46 | c.11614G>C | p.(Glu3872Gln) | Both | Likely Pathogenic | Conflicting classifications of pathogenicity | PS4-Mod,PM2-Sup,PM6,PP1-Sup,PP3,PP4 | LP |
| 42/46 | c.11657G>C | p.(Arg3886Pro) | ClinVar | - | Likely pathogenic | PS4-Sup,PM2-Sup,PP1-Str,PP4 | LP |
| 46/46 | c.12448C>T | p.(Arg4150Cys) | Both | Likely Pathogenic | Conflicting classifications of pathogenicity | PS3-Mod,PS4-Str,PM2-Sup,PP1-Sup,PP3,PP4 | P |

*^a^PKD1 NM_001009944.3*

*^b^PKD1 NP_001009944.3*

*Supplementary Table 2. Missense variants in PKD1 classified as pathogenic or likely pathogenic by ACMG/AMP criteria, including the reported classification in the PKDB/ClinVar databases and the criteria levels applied for each variant. P = Pathogenic; LP = Likely Pathogenic.*

| **Missense Variant*** | **SIFT**  **Score** | **PolyPhen Score** | **CADD Score** | **REVEL Score** | **AlphaMissense Score** |
| --- | --- | --- | --- | --- | --- |
| p.(Trp139Cys) | 0.01 | 0 | 23.7 | 0.846 | 0.9691 |
| p.(Cys210Gly) | 0 | 0 | 22.1 | 0.708 | 0.7525 |
| p.(Cys508Arg) | 0 | 1 | 25.7 | 0.759 | 0.9819 |
| p.(Arg510Pro) | 0 | 0.963 | 26.9 | 0.238 | 0.9782 |
| p.(Arg611Trp) | 0 | 0.898 | 25.7 | 0.324 | 0.4177 |
| p.(Ser699Pro) | 0.03 | 0.456 | 23.2 | 0.108 | 0.4501 |
| p.(Leu727Pro) | 0 | 0.991 | 23.8 | 0.422 | 0.5649 |
| p.(Leu845Ser) | 0 | 0.993 | 25.4 | 0.534 | 0.7967 |
| p.(Tyr1181Cys) | 0 | 0.992 | 19.75 | 0.819 | 0.6012 |
| p.(Val1206Gly) | 0.02 | 0.6 | 21.5 | 0.470 | 0.3361 |
| p.(Trp1414Ser) | 0 | 1 | 26.2 | 0.790 | 0.7906 |
| p.(Trp2298Gly) | 0 | 1 | 28.2 | 0.941 | 0.9016 |
| p.(Ser2372Ala) | 0 | 0.996 | 26.2 | 0.687 | 0.5193 |
| p.(Gly2391Asp) | 0 | 1 | 26.9 | 0.811 | 0.9367 |
| p.(Thr2422Lys) | 0 | 1 | 25.5 | 0.736 | 0.9459 |
| p.(Ser2423Phe) | 0 | 0.995 | 25.4 | 0.665 | 0.673 |
| p.(Leu2431Gln) | 0 | 1 | 26.2 | 0.911 | 0.9544 |
| p.(Arg2434Trp) | 0 | 1 | 31 | 0.754 | 0.2845 |
| p.(Cys2495Arg) | 0 | 1 | 23.0 | 0.875 | 0.9922 |
| p.(Arg2516Cys) | 0 | 1 | 33 | 0.793 | 0.6796 |
| p.(Arg2643Cys) | 0 | 1 | 32 | 0.693 | 0.366 |
| p.(Arg2767Cys) | 0 | 1 | 24.1 | 0.561 | 0.6322 |
| p.(Arg2767Pro) | 0 | 0.999 | 23.1 | 0.592 | 0.9408 |
| p.(Asn2770Ile) | 0 | 0.999 | 22.0 | 0.372 | 0.8981 |
| p.(Glu2771Lys) | 0 | 0.998 | 23.7 | 0.521 | 0.7987 |
| p.(Leu2816Pro) | 0 | 0.912 | 23.1 | 0.466 | 0.3896 |
| p.(His2921Pro) | 0 | 0.902 | 21.3 | 0.690 | 0.9709 |
| p.(Leu2993Pro) | 0 | 0.98 | 25.6 | 0.670 | 0.9752 |
| p.(Thr3126Ile) | 0 | 1 | 32 | 0.927 | 0.9854 |
| p.(Thr3135Met) | 0.01 | 1 | 26.3 | 0.844 | 0.4755 |
| p.(Phe3168Leu) | 0 | 0.999 | 23.7 | 0.716 | 0.9929 |
| p.(Lys3181Asn) | 0 | 0.997 | 20.1 | 0.372 | 0.8848 |
| p.(Pro3193Leu) | 0 | 1 | 22.2 | 0.705 | 0.9023 |
| p.(Arg3277Cys) | 0 | 1 | 23.6 | 0.634 | 0.5469 |
| p.(Gly3651Ser) | 0 | 1 | 28.5 | 0.298 | 0.6766 |
| p.(Arg3719Gln) | 0 | 0.981 | 34 | 0.505 | 0.1197 |
| p.(Trp3726Arg) | 0 | 0.49 | 26.4 | 0.726 | 0.9749 |
| p.(Arg3753Trp) | 0 | 0.994 | 33 | 0.873 | 0.4527 |
| p.(Leu3852Pro) | 0 | 0.834 | 28.8 | 0.653 | 0.7145 |
| p.(Glu3872Gln) | 0 | 1 | 24.9 | 0.834 | 0.6032 |
| p.(Arg3886Pro) | 0.04 | 0.831 | 19.16 | 0.514 | 0.5165 |
| p.(Arg4150Cys) | 0 | 1 | 34 | 0.676 | 0.7 |

**PKD1 NP_001009944.3*

*Supplementary Table 3. Missense variants in PKD1 classified as pathogenic or likely pathogenic by ACMG/AMP criteria, including output scores from in silico tools SIFT, PolyPhen, CADD, REVEL and AlphaMissense.*

| **cDNA**  **Change^a^** | **Missense**  **Variant^b^** | **ACMG Criteria Applied** | **ACMG Classifi-cation** | **SIFT**  **Score** | **PolyPhen Score** | **CADD Score** | **REVEL Score** | **AlphaMissense Score** |
| --- | --- | --- | --- | --- | --- | --- | --- | --- |
| c.82C>T | p.(Arg28Cys) | BP5, BP4, BS1 | LB | 0 | 0.685 | 25.6 | 0.045 | 0.6397 |
| c.107C>A | p.(Pro36His) | BP4, BS1 | LB | 0.08 | 0.047 | 21.3 | 0.083 | 0.2567 |
| c.169C>T | p.(Arg57Trp) | BP5, BP4, BS1 | LB | 0.01 | 0.042 | 22.3 | 0.134 | 0.1812 |
| c.182C>T | p.(Pro61Leu) | BS2_sup, BP4, BS1 | LB | 0.07 | 0.706 | 24.1 | 0.08 | 0.2374 |
| c.239G>A | p.(Arg80Gln) | BP5, BP4 | LB | 0.51 | 0.023 | 3.667 | 0.036 | 0.0656 |
| c.263C>T | p.(Ala88Val) | BS4, BP4, BS1 | B | 0.33 | 0.003 | 9.962 | 0.029 | 0.1051 |
| c.274G>A | p.(Ala92Thr) | BP5, BP4 | LB | 0.28 | 0.003 | 11.07 | 0.066 | 0.0842 |
| c.313A>T | p.(Thr105Ser) | BP5, BP4, BS1 | LB | 0.07 | 0.013 | 13.37 | 0.089 | 0.0971 |
| c.419C>T | p.(Ala140Val) | BP4, BS1 | LB | 1 | 0.003 | 0.097 | 0.268 | 0.069 |
| c.503G>C | p.(Gly168Ala) | BS1_sup, BP6, BP4 | LB | 0.04 | 0 | 12.48 | 0.031 | 0.0872 |
| c.580G>A | p.(Ala194Thr) | BP4, BS1 | LB | 0.09 | 0.017 | 12.85 | 0.006 | 0.0805 |
| c.603C>G | p.(His201Gln) | BP5, BP4, BS1 | LB | 0.22 | 0.003 | 0.030 | 0.09 | 0.1101 |
| c.739C>T | p.(Leu247Phe) | BP4, BS1 | LB | 1 | 0.015 | 0.963 | 0.055 | 0.0679 |
| c.827C>T | p.(Thr276Ile) | BP4, BS1 | LB | 0.03 | 0.087 | 20.4 | 0.068 | 0.1189 |
| c.871G>T | p.(Ala291Ser) | BP4, BS1 | LB | 0.65 | 0.017 | 5.758 | 0.105 | 0.0844 |
| c.971G>T | p.(Arg324Leu) | BP5, BS1 | LB | 0.04 | 0.344 | 16.68 | 0.358 | 0.1189 |
| c.1115G>A | p.(Ser372Asn) | BP5, BP4, BS1 | LB | 0.02 | 0.158 | 19.06 | 0.099 | 0.1564 |
| c.1117C>G | p.(Leu373Val) | BP5, BP4, BS1 | LB | 0.05 | 0.83 | 21.6 | 0.181 | 0.1202 |
| c.1202C>T | p.(Ala401Val) | BP4, BS1 | LB | 0.68 | 0 | 14.73 | 0.074 | 0.0564 |
| c.1354G>A | p.(Val452Met) | BP5, BP4 | LB | 0.02 | 0.794 | 17.72 | 0.038 | 0.1849 |
| c.1525G>A | p.(Val509Ile) | BP5, BP4 | LB | 0.02 | 0.999 | 23.9 | 0.175 | 0.1118 |
| c.1528C>T | p.(Arg510Trp) | BP5, BP4 | LB | 0 | 0.991 | 24.6 | 0.185 | 0.5344 |
| c.1594C>G | p.(Leu532Val) | BP5, BP4 | LB | 0.01 | 0.484 | 32 | 0.127 | 0.1639 |
| c.1621G>C | p.(Ala541Pro) | BP5, BP4 | LB | 0 | 0.922 | 21.9 | 0.135 | 0.514 |
| c.1666C>T | p.(Pro556Ser) | BP5, BP4 | LB | 0.1 | 0.335 | 17.07 | 0.108 | 0.0877 |
| c.1714C>T | p.(Pro572Ser) | BS2, BP4, BS1 | B | 0.28 | 0.059 | 13.23 | 0.017 | 0.0995 |
| c.1758A>C | p.(Glu586Asp) | BP4, BS1 | LB | 0.13 | 0.009 | 8.490 | 0.016 | 0.1616 |
| c.1801C>T | p.(Arg601Trp) | BP5, BP4, BS1 | LB | 0.02 | 0.64 | 18.95 | 0.053 | 0.098 |
| c.1870G>A | p.(Glu624Lys) | BS1_sup, BP6, BP4 | LB | 0.18 | 0.007 | 11.10 | 0.22 | 0.0727 |
| c.1885T>A | p.(Ser629Thr) | BP6, BP4 | LB | 0.35 | 0.012 | 2.346 | 0.143 | 0.0836 |
| c.1886C>T | p.(Ser629Phe) | BP4, BS1 | LB | 0.11 | 0.01 | 5.960 | 0.098 | 0.0845 |
| c.1910C>T | p.(Ala637Val) | BP5, BP4, BS1 | LB | 0.6 | 0.003 | 0.191 | 0.013 | 0.0758 |
| c.1922T>C | p.(Met641Thr) | BP5, BP4 | LB | 0.32 | 0.001 | 0.237 | 0.024 | 0.0798 |
| c.1942C>T | p.(Pro648Ser) | BP4, BS1 | LB | 0.12 | 0.516 | 21.3 | 0.101 | 0.0923 |
| c.1964C>T | p.(Pro655Leu) | BP5, BP4 | LB | 0.01 | 0.939 | 21.1 | 0.161 | 0.102 |
| c.1996G>A | p.(Ala666Thr) | BP5, BP4 | LB | 0.11 | 0.019 | 10.29 | 0.034 | 0.0816 |
| c.2039A>T | p.(Tyr680Phe) | BS2_sup, BP4, BS1 | LB | 0.04 | 0.89 | 16.89 | 0.108 | 0.1301 |
| c.2086G>A | p.(Ala696Thr) | BP5, BP4, BS1 | LB | 1 | 0.022 | 0.089 | 0.028 | 0.0701 |
| c.2143G>T | p.(Val715Phe) | BP5, BP4 | LB | 0.66 | 0.003 | 0.082 | 0.014 | 0.1922 |
| c.2176C>T | p.(Leu726Phe) | BP5, BP4, BS1 | LB | 0.75 | 0.023 | 0.127 | 0.025 | 0.0642 |
| c.2192C>T | p.(Pro731Leu) | BP4, BS1 | LB | 0.25 | 0.013 | 0.934 | 0.013 | 0.0671 |
| c.2222C>T | p.(Pro741Leu) | BP4, BS1 | LB | 0.48 | 0 | 3.086 | 0.013 | 0.086 |
| c.2227C>T | p.(Leu743Phe) | BS1_sup, BP4 | LB | 0.65 | 0 | 2.316 | 0.008 | 0.0909 |
| c.2239G>A | p.(Ala747Thr) | BS1_sup, BP6, BP4 | LB | 0.13 | 0.238 | 6.715 | 0.059 | 0.081 |
| c.2431C>G | p.(Leu811Val) | BP4, BS1 | LB | 0.05 | 0.153 | 10.09 | 0.097 | 0.0765 |
| c.2498A>G | p.(Asp833Gly) | BP5, BP4 | LB | 1 | 0.005 | 9.272 | 0.143 | 0.0828 |
| c.2504G>A | p.(Arg835His) | BP5, BP4 | LB | 0.72 | 0.003 | 0.025 | 0.036 | 0.0607 |
| c.2515C>T | p.(Pro839Ser) | BP5, BP4, BS1 | LB | 0.11 | 0.01 | 15.15 | 0.032 | 0.1147 |
| c.2527T>C | p.(Ser843Pro) | BS2, BP4, BS1 | B | 0.1 | 0.689 | 20.6 | 0.253 | 0.3128 |
| c.2548G>A | p.(Asp850Asn) | BP5, BP4, BS1 | LB | 0.25 | 0.337 | 19.24 | 0.096 | 0.1045 |
| c.2618T>C | p.(Val873Ala) | BP5, BP4, BS1 | LB | 0.97 | 0 | 12.45 | 0.018 | 0.0488 |
| c.2644G>A | p.(Val882Met) | BP5, BP4, BS1 | LB | 0.04 | 0.535 | 13.44 | 0.107 | 0.1147 |
| c.2707A>G | p.(Ser903Gly) | BP5, BP4, BS1 | LB | 1 | 0 | 0.016 | 0.064 | 0.0657 |
| c.2719C>T | p.(His907Tyr) | BP5, BP4 | LB | 0.63 | 0.001 | 14.81 | 0.128 | 0.0967 |
| c.2755C>T | p.(Arg919Trp) | BP5, BP4 | LB | 0.01 | 0.608 | 22.7 | 0.241 | 0.1121 |
| c.3101A>G | p.(Asn1034Ser) | BP5, BP4, BS1 | LB | 0.03 | 0.708 | 20.8 | 0.268 | 0.0837 |
| c.3101A>T | p.(Asn1034Ile) | BP5, BS1 | LB | 0 | 0.941 | 22.6 | 0.356 | 0.3908 |
| c.3113C>A | p.(Ala1038Glu) | BS1_sup, BP4 | LB | 1 | 0.003 | 0.004 | 0.185 | 0.1058 |
| c.3140C>T | p.(Ser1047Leu) | BP5, BS1 | LB | 0.01 | 0.913 | 26.1 | 0.419 | 0.1835 |
| c.3275T>C | p.(Met1092Thr) | BP4, BS1 | LB | 1 | 0.003 | 12.28 | 0.152 | 0.0374 |
| c.3277C>T | p.(His1093Tyr) | BP5, BS1 | LB | 0.36 | 0.985 | 20.7 | 0.429 | 0.1783 |
| c.3304C>G | p.(Leu1102Val) | BP4, BS1 | LB | 0.66 | 0.018 | 13.71 | 0.168 | 0.0635 |
| c.3316C>G | p.(Leu1106Val) | BP5, BP4, BS1 | LB | 0.71 | 0.102 | 1.437 | 0.057 | 0.0776 |
| c.3380C>T | p.(Pro1127Leu) | BP5, BS1 | LB | 0.18 | 0.368 | 16.60 | 0.37 | 0.11 |
| c.3406G>T | p.(Gly1136Cys) | BP5, BP4 | LB | 0.03 | 0.031 | 1.770 | 0.147 | 0.1045 |
| c.3425G>A | p.(Arg1142Gln) | BP6, BP4 | LB | 0.06 | 0.021 | 13.05 | 0.078 | 0.0841 |
| c.3424C>T | p.(Arg1142Trp) | BP5, BP4, BS1 | LB | 0.44 | 0.015 | 0.192 | 0.049 | 0.0676 |
| c.3430G>A | p.(Val1144Ile) | BP4, BS1 | LB | 0.28 | 0.062 | 0.599 | 0.1 | 0.0866 |
| c.3467G>C | p.(Gly1156Ala) | BP5, BS1 | LB | 0.01 | 0.957 | 18.57 | 0.292 | 0.1165 |
| c.3502C>T | p.(Pro1168Ser) | BP4, BS1 | LB | 1 | 0.249 | 7.292 | 0.068 | 0.0891 |
| c.3545C>T | p.(Ala1182Val) | BS1_sup, BP4 | LB | 0.17 | 0.035 | 1.578 | 0.051 | 0.0809 |
| c.3611C>T | p.(Ala1204Val) | BP4, BS1 | LB | 1 | 0.009 | 1.464 | 0.065 | 0.1492 |
| c.3611C>A | p.(Ala1204Glu) | BP4, BS1 | LB | 0.36 | 0.005 | 7.942 | 0.062 | 0.0779 |
| c.3679G>T | p.(Ala1227Ser) | BP4, BS1 | LB | 0.03 | 0.146 | 12.58 | 0.047 | 0.0838 |
| c.3868C>G | p.(Leu1290Val) | BP4, BS1 | LB | 0.04 | 0.059 | 15.75 | 0.144 | 0.0741 |
| c.3931G>A | p.(Ala1311Thr) | BP4, BS1 | LB | 0.1 | 0.052 | 16.50 | 0.106 | 0.0867 |
| c.3935G>A | p.(Arg1312Gln) | BP4, BS1 | LB | 1 | 0.01 | 0.012 | 0.041 | 0.0684 |
| c.3994G>A | p.(Asp1332Asn) | BS1 | LB | 0.05 | 0.953 | 24.7 | 0.586 | 0.3882 |
| c.4015G>A | p.(Val1339Met) | BP5, BP4, BS1 | LB | 0.02 | 0.878 | 11.63 | 0.13 | 0.1376 |
| c.4019G>A | p.(Arg1340Gln) | BP4, BS1 | LB | 0.18 | 0.021 | 5.135 | 0.293 | 0.0659 |
| c.4018C>T | p.(Arg1340Trp) | BP5, BS1 | LB | 0.34 | 0.01 | 0.010 | 0.077 | 0.0648 |
| c.4051C>T | p.(Arg1351Trp) | BP5, BP4, BS1 | LB | 0.01 | 0.929 | 20.6 | 0.135 | 0.0991 |
| c.4120A>T | p.(Ile1374Phe) | BP5, BP4 | LB | 0.02 | 0.547 | 17.94 | 0.259 | 0.2438 |
| c.4195T>C | p.(Trp1399Arg) | BP4, BS1 | LB | 1 | 0 | 4.907 | 0.091 | 0.0444 |
| c.4264G>A | p.(Ala1422Thr) | BP5, BP4, BS1 | LB | 0.36 | 0.015 | 0.132 | 0.018 | 0.0679 |
| c.4340C>T | p.(Ala1447Val) | BS2_sup, BP4, BS1 | LB | 1 | 0.014 | 0.107 | 0.168 | 0.0729 |
| c.4546G>A | p.(Ala1516Thr) | BP4, BS1 | LB | 0.28 | 0.015 | 0.003 | 0.017 | 0.07 |
| c.4669C>T | p.(Arg1557Cys) | BS2, BS1 | B | 0.03 | 0.929 | 25.8 | 0.312 | 0.1985 |
| c.4789A>G | p.(Ile1597Val) | BP5, BP4 | LB | 0 | 0.651 | 23.5 | 0.218 | 0.1312 |
| c.4810G>A | p.(Val1604Met) | BP5, BP4, BS1 | LB | 0 | 0.999 | 23.8 | 0.28 | 0.211 |
| c.4856C>T | p.(Ser1619Phe) | BP5, BP4, BS1 | LB | 0.02 | 0.909 | 20.2 | 0.259 | 0.1357 |
| c.4876G>A | p.(Val1626Ile) | BP4, BS1 | LB | 1 | 0.005 | 0.038 | 0.13 | 0.06 |
| c.4946C>T | p.(Thr1649Met) | BP5, BS1 | LB | 0.02 | 0.97 | 25.2 | 0.366 | 0.1358 |
| c.4963G>T | p.(Val1655Leu) | BP4, BS1 | LB | 0.05 | 0.013 | 0.177 | 0.111 | 0.1005 |
| c.5021C>T | p.(Pro1674Leu) | BP4, BS1 | LB | 0.24 | 0.006 | 3.133 | 0.086 | 0.0726 |
| c.5032G>A | p.(Gly1678Ser) | BP4, BS1 | LB | 0.44 | 0.035 | 11.66 | 0.111 | 0.0801 |
| c.5051C>T | p.(Ser1684Leu) | BP5, BP4, BS1 | LB | 0.1 | 0.021 | 10.21 | 0.092 | 0.0701 |
| c.5081A>G | p.(His1694Arg) | BS1_sup, BP4 | LB | 0.38 | 0.09 | 1.507 | 0.133 | 0.0624 |
| c.5318C>T | p.(Thr1773Ile) | BP5, BP4, BS1 | LB | 0.02 | 0.415 | 0.346 | 0.11 | 0.0923 |
| c.5357C>T | p.(Pro1786Leu) | BS4, BP5, BP4, BS1 | B | 0.01 | 0.224 | 13.39 | 0.15 | 0.072 |
| c.5363G>A | p.(Gly1788Asp) | BP6, BS1 | LB | 0.19 | 0.356 | 17.49 | 0.475 | 0.2482 |
| c.5369C>T | p.(Ala1790Val) | BP6, BS1_sup, BP4 | LB | 0.04 | 0.738 | 17.13 | 0.231 | 0.542 |
| c.5368G>C | p.(Ala1790Pro) | BS2, BP4 | LB | 0.48 | 0.009 | 0.349 | 0.128 | 0.0842 |
| c.5374G>A | p.(Ala1792Thr) | BS2, BP4 | LB | 0.11 | 0.174 | 10.80 | 0.209 | 0.08 |
| c.5418C>A | p.(Ser1806Arg) | BP5, BP4, BS1 | LB | 0.27 | 0.017 | 4.043 | 0.044 | 0.2438 |
| c.5433G>T | p.(Glu1811Asp) | BP5, BP4, BS1 | LB | 0.14 | 0.014 | 0.278 | 0.024 | 0.1046 |
| c.5458G>A | p.(Gly1820Arg) | BP5, BS1 | LB | 0 | 0.999 | 22.3 | 0.469 | 0.2129 |
| c.5582C>T | p.(Thr1861Ile) | BP5, BS1_sup | LB | 0.43 | 0.101 | 1.778 | 0.367 | 0.087 |
| c.5611G>A | p.(Ala1871Thr) | BA1 | B | 0.04 | 0.965 | 19.41 | 0.303 | 0.0912 |
| c.5633C>T | p.(Thr1878Met) | BS2_sup, BP4, BS1 | LB | 0.18 | 0.017 | 0.464 | 0.133 | 0.0863 |
| c.5662G>A | p.(Val1888Met) | BP4, BS1 | LB | 0.05 | 0.017 | 7.846 | 0.084 | 0.0898 |
| c.5777C>T | p.(Ala1926Val) | BP5, BP4 | LB | 0.05 | 0.224 | 9.327 | 0.091 | 0.087 |
| c.5783C>G | p.(Pro1928Arg) | BP5, BP4, BS1 | LB | 0.01 | 0.501 | 12.60 | 0.183 | 0.0921 |
| c.5825G>A | p.(Arg1942His) | BP5, BP4 | LB | 0.11 | 0.115 | 2.743 | 0.098 | 0.0779 |
| c.5827G>A | p.(Val1943Ile) | BP4, BS1 | LB | 0.17 | 0.502 | 13.04 | 0.111 | 0.1025 |
| c.5984G>A | p.(Arg1995His) | BS4, BP4, BS1 | B | 0.14 | 0.97 | 23.2 | 0.232 | 0.0944 |
| c.6001C>T | p.(Arg2001Trp) | BP5, BS1 | LB | 0.01 | 0.952 | 26.4 | 0.443 | 0.1641 |
| c.6007G>A | p.(Ala2003Thr) | BP5, BP4, BS1 | LB | 0.47 | 0.224 | 19.53 | 0.076 | 0.1161 |
| c.6151C>T | p.(Arg2051Cys) | BP4, BS1 | LB | 0.06 | 0.013 | 12.28 | 0.048 | 0.1118 |
| c.6185A>T | p.(Gln2062Leu) | BP5, BP4, BS1 | LB | 0.45 | 0.069 | 9.012 | 0.274 | 0.1432 |
| c.6200A>G | p.(Gln2067Arg) | BP5, BP4 | LB | 0.88 | 0.006 | 0.006 | 0.051 | 0.0556 |
| c.6205G>A | p.(Gly2069Ser) | BP4, BS1 | LB | 1 | 0.062 | 0.448 | 0.027 | 0.0906 |
| c.6224G>A | p.(Arg2075His) | BP4, BS1 | LB | 0.01 | 0.215 | 23.4 | 0.156 | 0.0872 |
| c.6244G>A | p.(Ala2082Thr) | BP5, BP4, BS1 | LB | 0.14 | 0.085 | 1.485 | 0.011 | 0.0714 |
| c.6299C>T | p.(Ser2100Leu) | BP5, BS1 | LB | 0 | 0.694 | 21.5 | 0.376 | 0.0981 |
| c.6331G>A | p.(Glu2111Lys) | BS2, BP4, BS1 | B | 0.13 | 0.29 | 12.07 | 0.078 | 0.1101 |
| c.6395T>G | p.(Phe2132Cys) | BP5, BS1 | LB | 0.01 | 0.993 | 23.8 | 0.293 | 0.545 |
| c.6439C>T | p.(Arg2147Trp) | BP4, BS1 | LB | 0 | 0.846 | 21.4 | 0.105 | 0.1418 |
| c.6457G>A | p.(Val2153Met) | BP5, BP4, BS1 | LB | 0.08 | 0.947 | 21.3 | 0.123 | 0.126 |
| c.6484C>T | p.(Arg2162Trp) | BP5, BP4, BS1 | LB | 0.35 | 0.018 | 21.9 | 0.164 | 0.187 |
| c.6488G>A | p.(Arg2163Gln) | BP5, BP4, BS1 | LB | 0 | 0.999 | 28.7 | 0.274 | 0.3192 |
| c.6545A>G | p.(Gln2182Arg) | BP5, BP4, BS1 | LB | 0.1 | 0.503 | 21.1 | 0.233 | 0.0754 |
| c.6572G>A | p.(Arg2191His) | BP5, BS1 | LB | 0.02 | 0.987 | 24.9 | 0.49 | 0.1161 |
| c.6593C>T | p.(Pro2198Leu) | BP5, BP4, BS1 | LB | 0.97 | 0.007 | 2.162 | 0.115 | 0.075 |
| c.6598C>T | p.(Arg2200Cys) | BP5, BP4, BS1 | LB | 0.02 | 0.86 | 22.8 | 0.233 | 0.2165 |
| c.6608G>A | p.(Arg2203His) | BS1_sup, BP4 | LB | 0.05 | 0.015 | 4.024 | 0.081 | 0.0957 |
| c.6625G>A | p.(Val2209Met) | BP5, BP4 | LB | 0.01 | 1 | 24.4 | 0.28 | 0.2085 |
| c.6635G>A | p.(Ser2212Asn) | BS1_sup, BP4 | LB | 0.59 | 0.186 | 16.16 | 0.113 | 0.1541 |
| c.6644G>A | p.(Arg2215Gln) | BP4, BS1 | LB | 1 | 0.003 | 19.22 | 0.173 | 0.0507 |
| c.6665C>T | p.(Ala2222Val) | BP5, BP4, BS1 | LB | 0.1 | 0.087 | 16.46 | 0.101 | 0.0984 |
| c.6749C>T | p.(Thr2250Met) | BP5, BS1 | LB | 0 | 0.995 | 25.6 | 0.579 | 0.2092 |
| c.6799G>A | p.(Val2267Met) | BP5, BS1 | LB | 0 | 1 | 24.8 | 0.411 | 0.2866 |
| c.6815G>A | p.(Arg2272Gln) | BS1_sup, BP4 | LB | 1 | 0.018 | 16.07 | 0.052 | 0.0546 |
| c.6878C>T | p.(Pro2293Leu) | BP6, BS1 | LB | 0.37 | 0.06 | 18.66 | 0.415 | 0.1071 |
| c.6905C>G | p.(Ala2302Gly) | BP5, BP4 | LB | 0.03 | 0.031 | 23.8 | 0.065 | 0.1068 |
| c.6928G>A | p.(Gly2310Arg) | BP5, BS1 | LB | 0.03 | 0.894 | 20.7 | 0.442 | 0.1101 |
| c.6935C>T | p.(Ala2312Val) | BP4, BS1 | LB | 0.37 | 0.015 | 1.428 | 0.143 | 0.0708 |
| c.6953G>A | p.(Arg2318His) | BP6, BS1 | LB | 0.03 | 0.942 | 25.3 | 0.374 | 0.1326 |
| c.6979C>T | p.(Arg2327Trp) | BP5, BS1 | LB | 0.04 | 0.86 | 22.7 | 0.325 | 0.182 |
| c.6986G>A | p.(Arg2329Gln) | BP5, BS1 | LB | 0.44 | 0.825 | 22.9 | 0.356 | 0.0815 |
| c.7124C>T | p.(Ala2375Val) | BS2, BS1 | B | 0 | 0.99 | 32 | 0.428 | 0.1962 |
| c.7147C>T | p.(Arg2383Cys) | BP5, BS1_sup | LB | 0 | 0.946 | 32 | 0.479 | 0.2728 |
| c.7190G>A | p.(Ser2397Asn) | BP4, BS1 | LB | 0.13 | 0.261 | 18.24 | 0.063 | 0.1059 |
| c.7210C>T | p.(Arg2404Trp) | BP5, BS1 | LB | 0 | 0.969 | 26.6 | 0.497 | 0.1198 |
| c.7241C>T | p.(Thr2414Met) | BP5, BS1 | LB | 0.01 | 0.996 | 25.8 | 0.505 | 0.1212 |
| c.7280C>T | p.(Ala2427Val) | BP5, BP4, BS1 | LB | 0.04 | 0.179 | 18.61 | 0.176 | 0.0967 |
| c.7324G>C | p.(Glu2442Gln) | BP5, BP6 | LB | 0.03 | 0.868 | 25.7 | 0.325 | 0.1554 |
| c.7391G>A | p.(Arg2464His) | BP4, BS1 | LB | 0.07 | 0.928 | 22.0 | 0.206 | 0.0642 |
| c.7430G>A | p.(Arg2477His) | BP6, BS1 | LB | 0.03 | 0.908 | 25.1 | 0.496 | 0.1492 |
| c.7429C>T | p.(Arg2477Cys) | BS4, BS1 | B | 0.24 | 0.007 | 12.09 | 0.362 | 0.0659 |
| c.7448C>T | p.(Ala2483Val) | BS1 | LB | 0.18 | 0.432 | 6.450 | 0.367 | 0.0857 |
| c.7463C>T | p.(Thr2488Ile) | BP5, BS1 | LB | 0.1 | 0.726 | 21.2 | 0.393 | 0.244 |
| c.7516G>A | p.(Ala2506Thr) | BP4, BS1 | LB | 0.38 | 0.159 | 18.07 | 0.13 | 0.0779 |
| c.7637A>G | p.(His2546Arg) | BP5, BP4 | LB | 0.07 | 0.471 | 16.65 | 0.381 | 0.0759 |
| c.7636C>T | p.(His2546Tyr) | BP5, BS1 | LB | 0.32 | 0.01 | 1.111 | 0.116 | 0.0565 |
| c.7642G>C | p.(Glu2548Gln) | BP4, BS1 | LB | 0.68 | 0.017 | 3.467 | 0.06 | 0.0759 |
| c.7712C>T | p.(Ala2571Val) | BS1_sup, BP4 | LB | 0.36 | 0.027 | 7.606 | 0.066 | 0.0822 |
| c.7745C>T | p.(Thr2582Met) | BP4, BS1 | LB | 0.21 | 0.03 | 0.075 | 0.143 | 0.0743 |
| c.7789C>G | p.(Pro2597Ala) | BP4, BS1 | LB | 0.05 | 0.508 | 21.5 | 0.172 | 0.1006 |
| c.7813C>T | p.(Pro2605Ser) | BP5, BS1_sup | LB | 0.19 | 0.739 | 22.5 | 0.414 | 0.2762 |
| c.7882G>A | p.(Val2628Met) | BP5, BP4 | LB | 0.3 | 0.007 | 0.450 | 0.012 | 0.0826 |
| c.7913A>G | p.(His2638Arg) | BP4, BS1 | LB | 0.08 | 0 | 12.17 | 0.1 | 0.0593 |
| c.7918G>C | p.(Ala2640Pro) | BS2, BP4 | LB | 0.02 | 0.766 | 24.6 | 0.209 | 0.36 |
| c.7960A>G | p.(Arg2654Gly) | BP5, BP4 | LB | 0.07 | 0.384 | 16.82 | 0.191 | 0.0899 |
| c.7993G>A | p.(Ala2665Thr) | BP4, BS1 | LB | 0.19 | 0.022 | 14.49 | 0.063 | 0.0698 |
| c.8020C>T | p.(Pro2674Ser) | BP4, BS1 | LB | 0.42 | 0 | 9.456 | 0.05 | 0.0808 |
| c.8087T>G | p.(Leu2696Arg) | BP5, BP4, BS1 | LB | 0.23 | 0.129 | 8.658 | 0.138 | 0.1789 |
| c.8087T>C | p.(Leu2696Pro) | BP5, BP4, BS1 | LB | 0.67 | 0 | 4.699 | 0.25 | 0.0426 |
| c.8111C>T | p.(Ala2704Val) | BP5, BP4, BS1 | LB | 0 | 0.082 | 14.02 | 0.117 | 0.103 |
| c.8123C>T | p.(Thr2708Met) | BP5, BS1 | LB | 0 | 0.999 | 22.1 | 0.392 | 0.2908 |
| c.8200C>A | p.(Pro2734Thr) | BP5, BP4, BS1 | LB | 0.13 | 0.082 | 5.177 | 0.011 | 0.0651 |
| c.8204A>T | p.(Gln2735Leu) | BP5, BP4, BS1 | LB | 0.02 | 0.052 | 14.32 | 0.099 | 0.0746 |
| c.8224G>A | p.(Glu2742Lys) | BP5, BP4, BS1 | LB | 0.11 | 0.017 | 12.39 | 0.072 | 0.0812 |
| c.8262C>A | p.(Asn2754Lys) | BS1_sup, BP4 | LB | 0.02 | 0.17 | 18.91 | 0.05 | 0.1969 |
| c.8282G>A | p.(Arg2761His) | BP4, BS1 | LB | 0.01 | 0.852 | 22.9 | 0.137 | 0.079 |
| c.8335G>A | p.(Glu2779Lys) | BP5, BP4, BS1 | LB | 0.1 | 0.541 | 18.44 | 0.188 | 0.0997 |
| c.8344G>A | p.(Val2782Met) | BS4, BP4, BS1 | LB | 0.13 | 0.416 | 12.77 | 0.079 | 0.0916 |
| c.8372G>A | p.(Arg2791Gln) | BS1 | LB | 0.03 | 0.608 | 21.2 | 0.11 | 0.0956 |
| c.8371C>T | p.(Arg2791Trp) | BP5, BP4 | LB | 0.57 | 0.006 | 5.520 | 0.294 | 0.0807 |
| c.8440G>A | p.(Gly2814Arg) | BP4, BS1 | LB | 0.05 | 0.582 | 20.9 | 0.165 | 0.1414 |
| c.8444C>T | p.(Ala2815Val) | BS2_sup, BP4, BS1 | LB | 0.12 | 0.026 | 16.01 | 0.054 | 0.1051 |
| c.8473C>T | p.(Leu2825Phe) | BP5, BP4 | LB | 0 | 0.861 | 21.2 | 0.15 | 0.3342 |
| c.8593C>T | p.(Arg2865Trp) | BP4, BS1 | LB | 0 | 0.715 | 23.3 | 0.285 | 0.1257 |
| c.8609G>A | p.(Arg2870His) | BP6, BS1 | LB | 0.05 | 0.791 | 22.8 | 0.376 | 0.1291 |
| c.8620G>A | p.(Val2874Met) | BP5, BS1 | LB | 0 | 0.999 | 21.9 | 0.594 | 0.5307 |
| c.8644T>A | p.(Trp2882Arg) | BP4, BS1 | LB | 0.57 | 0.033 | 7.128 | 0.076 | 0.0815 |
| c.8662C>G | p.(Arg2888Gly) | BP5, BP4 | LB | 0.32 | 0.001 | 6.690 | 0.007 | 0.0606 |
| c.8689G>A | p.(Val2897Ile) | BS4, BP4, BS1 | B | 0.52 | 0.119 | 1.977 | 0.063 | 0.1088 |
| c.8713G>A | p.(Val2905Ile) | BS2_sup, BP4, BS1 | LB | 0.05 | 0.082 | 16.67 | 0.098 | 0.1184 |
| c.8716G>A | p.(Gly2906Ser) | BP4, BS1 | LB | 1 | 0.001 | 0.931 | 0.051 | 0.0555 |
| c.8750C>T | p.(Ala2917Val) | BP4, BS1 | LB | 0.01 | 0.096 | 13.64 | 0.242 | 0.1149 |
| c.8780C>T | p.(Thr2927Met) | BP5, BP4, BS1 | LB | 0.03 | 0.909 | 18.18 | 0.28 | 0.1254 |
| c.8898G>C | p.(Glu2966Asp) | BP4, BS1 | LB | 0.41 | 0.006 | 2.638 | 0.029 | 0.1265 |
| c.8914G>A | p.(Asp2972Asn) | BP5, BP4, BS1 | LB | 0 | 0.984 | 24.5 | 0.237 | 0.1926 |
| c.8921G>A | p.(Arg2974Gln) | BP5, BP4, BS1 | LB | 0.21 | 0.95 | 22.2 | 0.147 | 0.1166 |
| c.8953A>G | p.(Arg2985Gly) | BP4, BS1 | LB | 0.59 | 0 | 0.006 | 0.159 | 0.0593 |
| c.9022G>A | p.(Val3008Met) | BS1 | LB | 0 | 0.981 | 23.9 | 0.316 | 0.2271 |
| c.9068T>A | p.(Met3023Lys) | BS1 | LB | 1 | 0.006 | 12.02 | 0.325 | 0.0773 |
| c.9154G>A | p.(Gly3052Ser) | BP5, BS1_sup | LB | 0 | 0.956 | 24.2 | 0.664 | 0.5468 |
| c.9169G>A | p.(Val3057Met) | BP5, BP4, BS1 | LB | 0 | 0.968 | 24.0 | 0.368 | 0.319 |
| c.9187C>T | p.(Arg3063Cys) | BP5, BP4, BS1 | LB | 0.01 | 0.535 | 20.9 | 0.151 | 0.1472 |
| c.9202G>C | p.(Glu3068Gln) | BP5, BP4 | LB | 0.15 | 0.072 | 25.2 | 0.105 | 0.0914 |
| c.9344G>A | p.(Arg3115Gln) | BP5, BP4, BS1 | LB | 0.02 | 0.129 | 21.0 | 0.121 | 0.0775 |
| c.9350G>A | p.(Arg3117His) | BP4, BS1 | LB | 0.05 | 0.608 | 24.4 | 0.28 | 0.113 |
| c.9349C>T | p.(Arg3117Cys) | BP5, BP4, BS1 | LB | 0.17 | 0.003 | 19.05 | 0.046 | 0.0848 |
| c.9421A>C | p.(Met3141Leu) | BS2_sup, BP4 | LB | 0.04 | 0.136 | 20.8 | 0.201 | 0.2417 |
| c.9506G>A | p.(Arg3169Gln) | BP4, BS1 | LB | 1 | 0.003 | 17.33 | 0.123 | 0.0581 |
| c.9548G>A | p.(Arg3183Gln) | BP5, BS1 | LB | 0 | 0.994 | 31 | 0.525 | 0.1962 |
| c.9620C>T | p.(Thr3207Met) | BP5, BS1 | LB | 0 | 0.975 | 13.69 | 0.29 | 0.1276 |
| c.9626G>A | p.(Arg3209His) | BP4, BS1 | LB | 0.02 | 0.279 | 21.0 | 0.346 | 0.1146 |
| c.9625C>T | p.(Arg3209Cys) | BP5, BS1 | LB | 0.04 | 0.026 | 16.32 | 0.163 | 0.0722 |
| c.9631G>A | p.(Ala3211Thr) | BP4, BS1 | LB | 0.39 | 0.012 | 13.02 | 0.119 | 0.0566 |
| c.9718G>A | p.(Ala3240Thr) | BP4, BS1 | LB | 0.45 | 0.039 | 11.70 | 0.078 | 0.0757 |
| c.9731G>A | p.(Arg3244His) | BP5, BP4, BS1 | LB | 0.23 | 0.579 | 16.41 | 0.1 | 0.0756 |
| c.9740G>A | p.(Arg3247His) | BP5, BS1 | LB | 0.02 | 0.999 | 28.2 | 0.334 | 0.1882 |
| c.9853G>A | p.(Val3285Ile) | BS1_sup, BP4 | LB | 0.46 | 0.003 | 4.510 | 0.018 | 0.0843 |
| c.9932A>G | p.(His3311Arg) | BP5, BP4, BS1 | LB | 0.1 | 0.013 | 4.869 | 0.099 | 0.0568 |
| c.10099A>G | p.(Ile3367Val) | BP5, BP4, BS1 | LB | 0.15 | 0.023 | 0.011 | 0.036 | 0.0703 |
| c.10102G>A | p.(Asp3368Asn) | BP4, BS1 | LB | 0.01 | 0.271 | 21.8 | 0.122 | 0.1 |
| c.10123G>A | p.(Val3375Met) | BP4, BS1 | LB | 0.45 | 0.029 | 12.55 | 0.15 | 0.0827 |
| c.10225G>C | p.(Val3409Leu) | BP4, BS1 | LB | 0.06 | 0.013 | 7.940 | 0.043 | 0.1332 |
| c.10234C>T | p.(Pro3412Ser) | BP5, BP4, BS1 | LB | 0.31 | 0.251 | 18.22 | 0.065 | 0.1241 |
| c.10240G>A | p.(Gly3414Ser) | BP5, BP4, BS1 | LB | 1 | 0 | 16.04 | 0.11 | 0.0548 |
| c.10304G>A | p.(Arg3435Gln) | BP4, BS1 | LB | 1 | 0 | 12.58 | 0.067 | 0.0625 |
| c.10325C>T | p.(Ala3442Val) | BP5, BP4, BS1 | LB | 0.09 | 0.052 | 3.911 | 0.043 | 0.0777 |
| c.10354G>A | p.(Gly3452Ser) | BS1_sup, BP4 | LB | 1 | 0.04 | 0.010 | 0.036 | 0.0649 |
| c.10387A>G | p.(Lys3463Glu) | BP4, BS1 | LB | 0 | 0.013 | 18.16 | 0.097 | 0.1267 |
| c.10400C>T | p.(Ala3467Val) | BP5, BP4, BS1 | LB | 0.02 | 0.015 | 17.88 | 0.047 | 0.0814 |
| c.10437G>C | p.(Glu3479Asp) | BP4, BS1 | LB | 1 | 0.003 | 0.630 | 0.065 | 0.0878 |
| c.10485C>A | p.(Asp3495Glu) | BP5, BP4 | LB | 0.03 | 0.964 | 18.87 | 0.125 | 0.1974 |
| c.10529C>T | p.(Thr3510Met) | BP5, BP4, BS1 | LB | 0.12 | 0.074 | 6.786 | 0.103 | 0.0738 |
| c.10531C>G | p.(Leu3511Val) | BP4, BS1 | LB | 0.43 | 0.019 | 0.379 | 0.029 | 0.0695 |
| c.10535C>T | p.(Ala3512Val) | BP4, BS1 | LB | 0.28 | 0.007 | 0.743 | 0.035 | 0.075 |
| c.10619G>C | p.(Gly3540Ala) | BP5, BP4, BS1 | LB | 1 | 0.622 | 22.2 | 0.123 | 0.0995 |
| c.10676A>C | p.(His3559Pro) | BP5, BP4 | LB | 0.01 | 0.859 | 23.6 | 0.269 | 0.2085 |
| c.10678G>A | p.(Gly3560Arg) | BP5, BS1 | LB | 0.05 | 0.973 | 23.0 | 0.352 | 0.7085 |
| c.10765C>T | p.(Leu3589Phe) | BP5, BS1 | LB | 0 | 0.995 | 22.8 | 0.435 | 0.265 |
| c.11015G>A | p.(Arg3672Gln) | BP5, BP4, BS1 | LB | 0.01 | 0.17 | 18.18 | 0.108 | 0.103 |
| c.11080T>C | p.(Cys3694Arg) | BP5, BP4 | LB | 1 | 0.007 | 8.747 | 0.283 | 0.1361 |
| c.11333C>A | p.(Thr3778Asn) | BP4, BS1 | LB | 0.03 | 0.179 | 21.5 | 0.126 | 0.2459 |
| c.11337C>G | p.(Ser3779Arg) | BP5, BP4, BS1 | LB | 0.14 | 0.03 | 0.023 | 0.16 | 0.1667 |
| c.11356G>C | p.(Glu3786Gln) | BP5, BP4, BS1 | LB | 0.28 | 0.03 | 12.31 | 0.042 | 0.071 |
| c.11520C>A | p.(His3840Gln) | BP5, BS1_sup | LB | 0.03 | 0.516 | 21.4 | 0.454 | 0.3292 |
| c.11627C>T | p.(Ala3876Val) | BP5, BS1 | LB | 0.03 | 0.75 | 21.6 | 0.311 | 0.1267 |
| c.11689C>T | p.(Leu3897Phe) | BP5, BS1 | LB | 0 | 0.965 | 23.5 | 0.339 | 0.1257 |
| c.11717G>T | p.(Cys3906Phe) | BP5, BS1 | LB | 1 | 0.003 | 7.426 | 0.632 | 0.1437 |
| c.11870G>A | p.(Gly3957Asp) | BP5, BS1 | LB | 0.08 | 0.51 | 16.14 | 0.441 | 0.3223 |
| c.11960C>G | p.(Ala3987Gly) | BS4, BP6 | LB | 0.01 | 0.792 | 24.6 | 0.305 | 0.1378 |
| c.12133A>G | p.(Ile4045Val) | BP4, BS1 | LB | 0.58 | 0.003 | 0.042 | 0.058 | 0.0714 |
| c.12176C>T | p.(Ala4059Val) | BP4, BS1 | LB | 0.08 | 0.015 | 7.495 | 0.091 | 0.0875 |
| c.12220C>A | p.(Leu4074Met) | BP5, BP4 | LB | 0.11 | 0.928 | 14.50 | 0.23 | 0.1359 |
| c.12292G>A | p.(Ala4098Thr) | BP5, BP4 | LB | 0.06 | 0.837 | 21.3 | 0.245 | 0.0955 |
| c.12370C>T | p.(Pro4124Ser) | BP5, BP4 | LB | 0.21 | 0.601 | 23.6 | 0.127 | 0.3269 |
| c.12436G>A | p.(Val4146Ile) | BP4, BS1 | LB | 0.08 | 0.642 | 23.6 | 0.072 | 0.2211 |
| c.12569C>T | p.(Ser4190Phe) | BS4, BP4, BS1 | LB | 0 | 0.999 | 24.5 | 0.175 | 0.1634 |
| c.12648A>C | p.(Gln4216His) | BP4, BS1 | LB | 0.17 | 0 | 0.024 | 0.289 | 0.0935 |
| c.12691C>A | p.(Gln4231Lys) | BS1, BP4 | LB | 0.09 | 0.535 | 22.7 | 0.045 | 0.1009 |
| c.12769G>A | p.(Gly4257Arg) | BP5, BP4, BS1 | LB | 0.27 | 0.001 | 7.727 | 0.012 | 0.1025 |
| c.12862A>G | p.(Ser4288Gly) | BP5, BP4 | LB | 0.27 | 0.054 | 0.297 | 0.022 | 0.0612 |

*^a^PKD1 NM_001009944.3*

*^b^PKD1 NP_001009944.3*

*Supplementary Table 4. Missense variants in PKD1 reported in PKDB as likely benign and classified as benign or likely benign by ACMG/AMP criteria, including the output scores from in silico tools SIFT, PolyPhen, CADD, REVEL and AlphaMissense.*

|  | **CADD** | **REVEL** | **AlphaMissense** |
| --- | --- | --- | --- |
| **AUC –**  **All Variants (n = 311)** | 0.895 | 0.956 | 0.990 |
| **AUC –**  **Only Variants Classified Independent of PP3/BP4 Criteria (n = 237)** | 0.882 | 0.924 | 0.992 |
| **Change in AUC** | -0.013 | -0.032 | +0.002 |
| **P-Value** | 0.69 | 0.29 | 0.78 |

*Supplementary Table 5. The Area Under the Curve (AUC) from the ROC curve analyses of the original set of variants, and after being re-run with the subset of variants with pathogenic or benign classification independent of PP3/BP4 evidence.*
